# Impacts of vaccination on the variant selection of SARS-CoV-2 and the subsequent case hospitalization rate: insights from the first U.S. Omicron wave

**DOI:** 10.1101/2024.10.03.24314829

**Authors:** Jingbo Liang, Zhaojun Ding, Qingpeng Zhang, Hsiang-Yu Yuan

**Affiliations:** Department of Biomedical Sciences, Jockey Club College of Veterinary Medicine and Life Sciences, City University of Hong Kong, Hong Kong SAR, China; Centre for Applied One Health Research and Policy Advice, Jockey Club College of Veterinary Medicine and Life Sciences, City University of Hong Kong, Hong Kong SAR, China; Musketeers Foundation Institute of Data Science, The University of Hong Kong, Hong Kong, China; Department of Pharmacology and Pharmacy, LKS Faculty of Medicine, The University of Hong Kong, Hong Kong, China

**Keywords:** COVID-19 vaccine, case hospitalization rate, viral evolution

## Abstract

**Background:** COVID-19 vaccines, while providing protection against hospitalization, could inadvertently increase selection pressure on new immune-escape variants, impacting case hospitalization rate (CHR).

**Methods:** Using epidemiological and genomic sequence data, we calculated daily state-level CHR and the proportion of Omicron mutations in the United States during the first Omicron wave (between December 11, 2021, and March 22, 2022). We derived mathematical formulas to link evolution responses to an increasing population immunity with lagged regression models. Using mediation analysis, together with generalized linear mixed models and distributed lag nonlinear models, we assessed how natural selection, shaped by vaccine coverage, impacts CHR.

**Results:** The model showed that increasing vaccination coverage from 45% to 70% contributed to a reduction in CHR from 5.8% to 4.4%. Part of the reduction resulted from direct vaccine protection (OR: 0.85, p-value=0.012). However, the higher vaccination coverage was correlated with a 20% increase in the proportion of BA.1/BA.1.1-associated mutations. As the Omicron variants were less severe than their predecessors (Delta), CHR further reduced (OR: 0.87, p-value<0.001). Marginally, this could reduce CHR from 5.8% to 5.1% via the adaptation of Omicron variants as marginal effect without accounting for direct vaccine protection.

**Conclusions:** The study offers new insight into vaccine strategies for reducing hospitalization risk by shortening [or maintaining] the circulation of more [or less] virulent variants among infectious diseases. Continuous monitoring of variant evolution, including their virulence, is critical.

## Introduction

Even though the global health emergency for the COVID-19 pandemic was declared over in May 2023 ^1^, concerns remain about the SARS-CoV-2’s ability to mutate and evade neutralizing antibodies ^2–4^. This emphasizes the critical need for continuous surveillance and forecasting of COVID-19 hospital admission rates to assess the demand on healthcare system against emerging variants of concern (VOC) ^5^, which have frequently emerged and swiftly become predominant since vaccine rollout. Besides vaccine effectiveness against severe illness or hospitalization ^6–8^, it is important to understand the impacts of vaccination on the adaptation of new immune-escape variants ^9^, which also influence the disease severity. Although in theory, selection pressures contribute to the fixation of beneficial mutations ^10,11^, evidence from observational studies using SARS-CoV-2 real-world data is lacking.

After the rollout of vaccines and the emergence of the Omicron variant, a significant decrease in case hospitalization rate (CHR) has been observed in many countries ^12,13^. The CHR, defined as the proportion of daily new hospital admissions relative to daily new reported cases, can be used as a metric to assess disease severity. This metric has been used to infer the number of hospitalizations in recent modelling studies to predict healthcare demand ^14^.

The observed reduction in CHR resulted from both vaccination and the replacement of the Delta strain with the less virulent (in terms of disease severity) ^15^. However, disentangling their effects is not straightforward, because of the complex interactions among vaccination, viral transmission, and viral adaptation ^11^.

In the United States (US), the first wave of Omicron (including BA1, BA1.1 and BA.2) caused over 27 million COVID-19 cases and about one million hospitalizations ^16^. Since the commencement of vaccination campaigns in December 2020, a distinct variation in vaccination coverage among states has been observed ^16^. However, the effects of these differences on CHR via the selection of the Omicron’s variants was largely unexplored.

Our study aimed to quantify the impact of vaccination on CHR through both the effectiveness against hospitalization and the selection of certain key mutations of variants and subvariants using causal mediation analysis. This helps to understand the relationship between vaccination, viral adaptation, and hospitalization. The findings provide insights into the complex role of vaccination in strengthening healthcare systems to effectively manage future outbreaks.

## Methods

### Data collection

This study focused on the first wave of the Omicron variant in the US, spanning from December 11, 2021, to March 22, 2022 with 27,256,687 COVID-19 cases and 980,036 COVID-19-associated hospitalizations. We defined the study period as beginning when the proportion of newly emerged Omicron variants exceeded 5% and ending when the daily reported cases of the first Omicron wave reached a minimum level preceding the subsequent wave (Figure S1). We collected daily state-level data on COVID-19 cases, hospital admissions, and vaccination rates from the Centers for Disease Control and Prevention ^16^.

The vaccine data included the daily cumulative percentage of fully vaccinated individuals (either with a second dose of a two-dose vaccine or one dose of a single-dose vaccine) (Figure S2), and daily cumulative percentage of people who received a booster.

We downloaded a total of 3,149,650 SARS-Cov-2 virus sequences from GISAID database ^17^, covering all 50 states of the US and the District of Columbia (Table S3). Our analysis focused on 19 key mutations on the Receptor Binding Domain (RBD) of the Omicron variant^18^, which were grouped into three categories: (i) BA.1/BA.1.1-associated mutations, including S371L, G496S, and G446S, (ii) shared mutations between BA.1/BA.1.1 and BA.2, including G339D, S373P, S375F, K417N, N440K, S477N, T478K, E484A, Q493R, Q498R, N501Y, and Y505H, and (iii) BA.2-associated mutations, including S371F, T376A, R408S, and D405N (Figure 2D and Figure S5). See “SARS-CoV-2 genomic surveillance data management” in Supplementary Materials for the procedure of the calculation of mutations.

Daily average temperature (Figure S3) and relative humidity (Figure S4) were collected from the National Center for Environmental Information ^18^. The number of staffed inpatient beds available each day was collected from the Department of Health and Human Services ^19^.

Daily average temperature and relative humidity for each state were computed by aggregating data from all monitoring stations within the respective state.

### Adjusted effective vaccination coverage considering boosting and waning immunities

Our analysis assumed that the effectiveness of the full vaccine against hospitalization decreased to its lowest level after six months due to waning immunity, remained stable thereafter, and then was restored after receiving a booster dose ^20^. Based on this assumption, we calculated the effective vaccination coverage (hereafter vaccination coverage, Vac_l_(t)) by summing the coverage after adjusting for vaccine waning and booster effects ^21^. Please see Supplementary Material for the detailed calculations.

### Mediation analysis

Causal mediation analysis ^22^ was performed to determine whether virus mutation mediated the relationship between vaccination coverage and CHR (Figure 1). This analysis followed the classical causal steps approach 22 in three steps: Step 1 (Basic model) involved modeling the total effect of vaccination coverage on CHR (i.e., path a in Figure 1); Step 2 (Intermediate model) involved modeling the effect of vaccination coverage on the proportion of virus mutations (i.e., path b); and Step 3 (Mediation model) involved modeling the direct effect (i.e., path a’) and indirect effect (i.e., path c) of vaccination coverage on CHR using. Here, the direct effect refers to vaccine effectiveness against hospitalization and the indirect effect refers to the change in CHR resulting from natural selection on immune-escape mutations. If all of these paths (a, b, c, and a’) are significant, and the direct effect is smaller than the total effect (a’ < a), the proportion of virus mutations can be concluded to be a mediator between vaccine and CHR.

**Figure 1.**
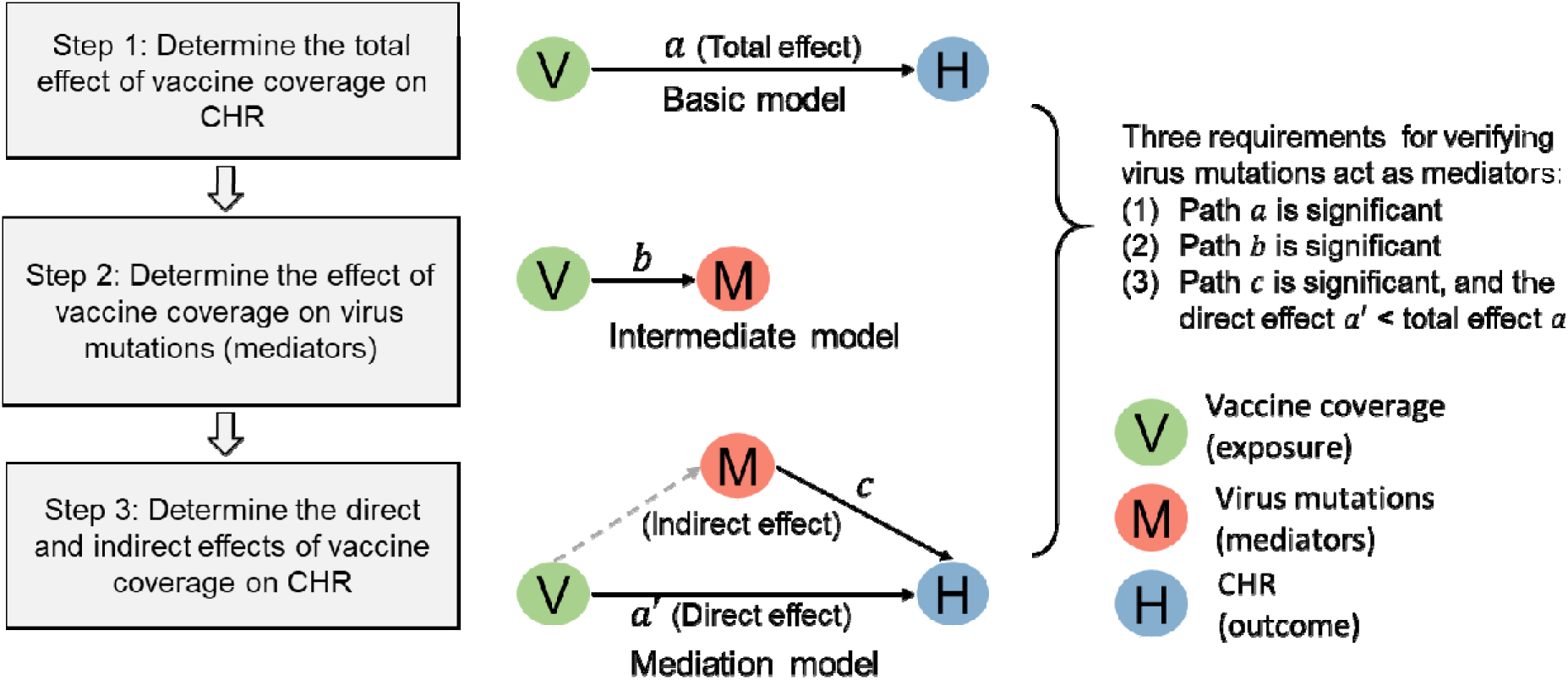
The flow of determining the mediation effect of Omicron mutations on the relationship between vaccination coverage and the COVID-19 CHR. Three steps were required to validate the mediation effect. Each step modelled the effect of paths using one of the three models (i.e., *basic (total effect)*, *intermediate,* and *mediation model*). Vaccination coverage (V) represented the exposure; virus mutations (M) represented the mediators; and CHR (H) represented the outcome in the models.

The *Intermediate model*, which investigated the impact of vaccination on the selection coefficient for the Omicron variant, was formulated based on the evolutionary process developed by Haldane JBS ^23^ and the selection coefficient developed by Chevin LM ^24^.

The selection coefficient, denoted by s, was defined as the rate of change in the logit-transformed frequency of a new variant over time: 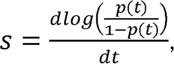, where is thproportion of the new variant at t, the time since the growth of the variant. If selection coefficient s was set to be proportional to the percentage of the hosts who are vaccinated (i.e. effective vaccine coverage *v_act_* as we used), we showed that the logit of new variant can be modelled as 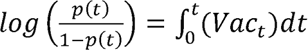 (see “Determine the effect of vaccination 1-p(t) o coverage on transmission of virus mutations (Intermediate Models)” in the Supplementary Material). Furthermore, in a discrete-time setting with nonlinear effects, 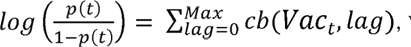, where *cb* is the crossbasis function and *Max* is the maximum considered lag time. To account for both state-specific effects and nonlinear vaccine lag effects, we utilized generalized linear mixed models (GLMM) integrated with distributed lag non-linear models (DLNM). Hence, the equations of the three models are described as follows:

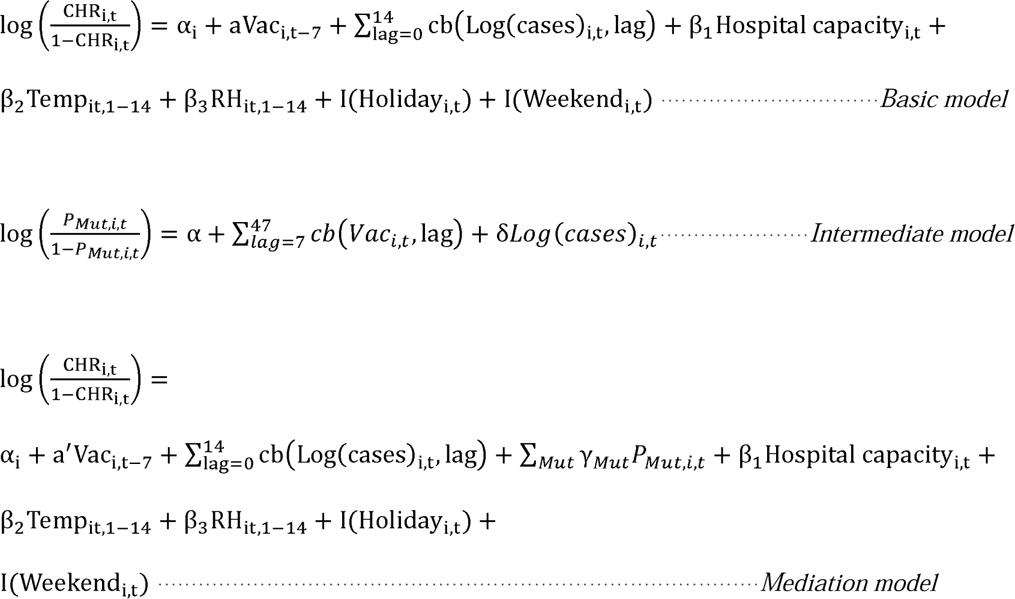

where *CHR_i,t_* represented the daily COVID-19 case hospitalization rate for state *i* on day *t*, and α*_i_* was the state-level random intercept in the basic and mediation model. *vac_i,t_* was the effective vaccination coverage, and ch(*vac_i,t_*, lag) was a cross-basis function that modeled the lag-response relationship between vaccination coverage and CHR. The vaccination lag time was set between 7 and 47 days, assuming vaccinations produce protection around one week after the shot. *P_Mut,i,t_* referred to *P_BA1,i,t_*, *P_BA12,i,t_*, and *P_BA2,i,t_*, the daily mean proportion of BA.1/BA.1.1-associated mutations, shared mutations between BA.1/BA.1.1 and BA.2 subvariants, and BA.2-associated mutations, respectively. The dependent variable is 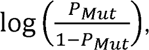, which has been used to estimate the selection coefficient on the vaccine escape mutations in a previous modelling study (see equation 1) 9.

### Potential confounders

Daily COVID-19 cases, hospital capacity, meteorological factors, and weekend/holiday indices, were added into the model according to the forward-stepwise AIC approach. The best-fitting model (i.e. with the smallest AIC) was chosen as the final Basic model (Table S4). Daily COVID-19 cases (i.e., *cases_i,t_*) were included with a logarithmically transformed relationship to CHR based on our observations (see Figure S13A). This accounted for the impact of public health interventions on the incidence of infections (healthcare system load) and therefore the CHR 25. Hospital capacity (*Hospital capacity_i,t_*) was measured by the daily number of staffed inpatient beds available. Meteorological conditions, known to be associated with COVID-19 severity 26, were accounted using 14-day moving averages of ambient temperature (*Temp_it,1-14_*) and relative humidity (*RH_it,1-14_*). Weekend and public holiday effects were indexed as I(*Weewend_it_*) and I(*Holiday_it_*). A detailed description of the models was provided in the Supplementary Material.

### Impact of prior infection and sensitivity analysis

To account for the immunity induced by prior natural infection, infection proportions were incorporated into the *Basic*, *Intermediate*, and *Mediation models* in our sensitivity analysis. The cumulative incidence of infections during the Delta wave (the latest before the study period) was added to the effective vaccination coverage to represent total population immunity. Infections prior to the Delta wave were excluded based on the assumption of significantly waned immunity.

Moreover, we conducted another sensitivity analysis to assess the impact of decreased vaccine effectiveness against hospitalization, assuming a decline to 80% efficacy after six months of full vaccination.

## Results

### Overview of the first Omicron wave in the United States

During the first Omicron wave, the number of daily reported cases and hospital admissions peaked in mid-January 2022, while the CHR experienced a temporary decline (Figure S1 and Figure S6). The vaccination coverage varied significantly among states, seemingly reducing cumulative hospitalizations per million population (Figure 2AB). For example, some Southern states such as Oklahoma (OK), Arkansas (AR), and Alabama (AL) had lower vaccination coverage (below 52%) concomitant with higher hospitalization burdens (exceeding 3,669 per million); Conversely, some Northeastern states like Vermont (VT) and Maine (ME) displayed higher vaccination coverage (above 64%) and lower hospitalization burdens. Concurrently, the Delta variant was replaced by the Omicron variant (BA.1) and subsequently by its subvariants, mainly BA 1.1 and BA. 2 (Figure 2C).

**Figure 2.**
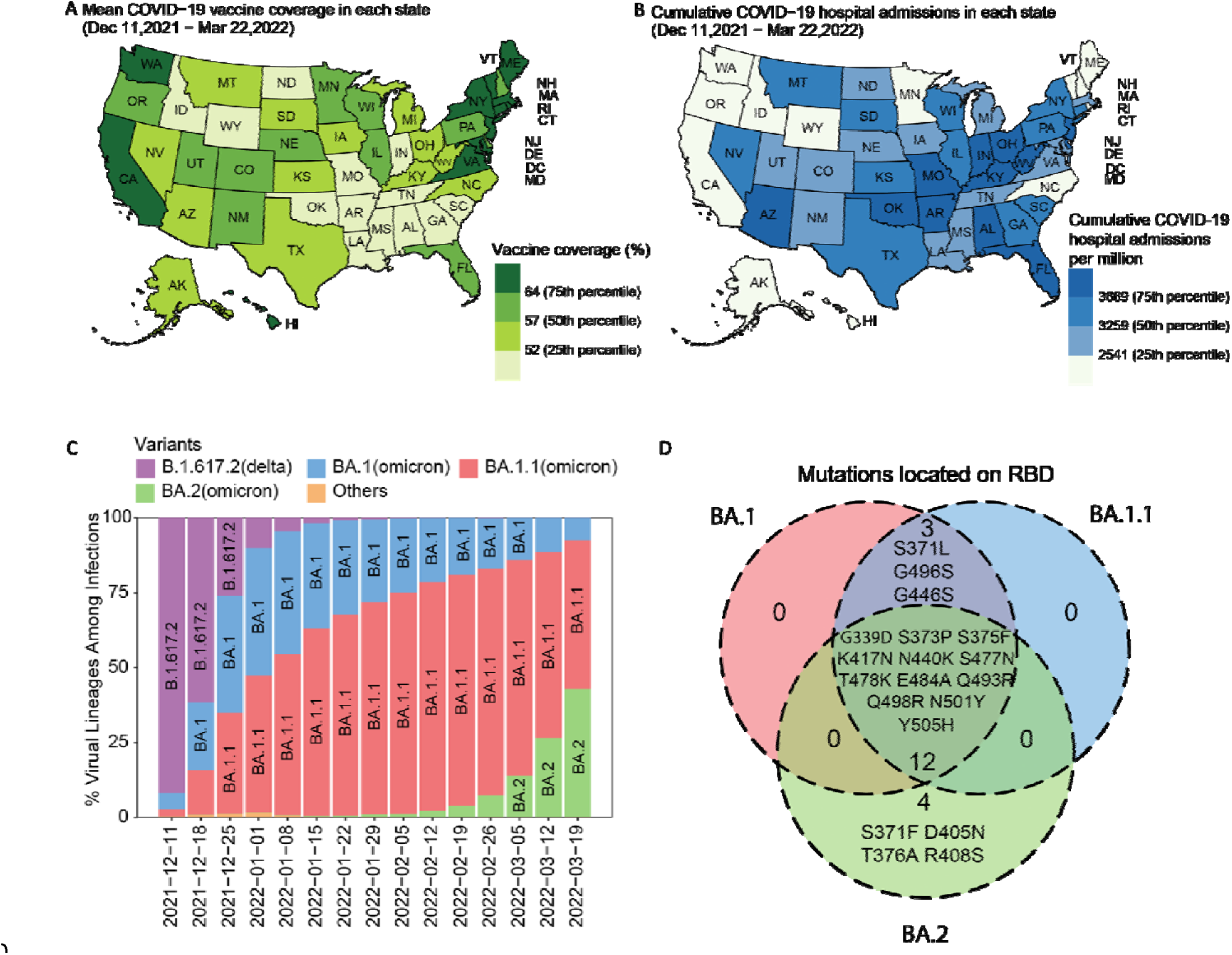
Vaccination coverages, cumulative hospital admissions, and the proportion of virus variants during the first Omicron wave in the United States. (A) Average vaccination coverage during the first Omicron wave (i.e., December 11, 2021 to March 22, 2022) at the state level. (B) The state-level cumulative COVID-19 hospital admissions (per million) during the first Omicron wave. (C) The proportion of variants that circulated in the US during the first Omicron wave (obtained from CDC ^14^). (D) Venn plots of the important mutations (RBD) of the BA.1, BA.1.1, and BA.2 subvariants.

### State-level analysis of transmission and CHR

We conducted a comparison of transmission and CHR between two groups of states: states with high vaccination coverage (Group H) and states with low vaccination coverage (Group L). The classification was based on whether their average coverage during the first Omicron wave was above or below the national average of 59% across all states (see Table S1 for the list of states by group). No significant difference was found in infection incidences between Groups H and L (p-value > 0.05, Figures 3AC and S7A).

**Figure 3.**
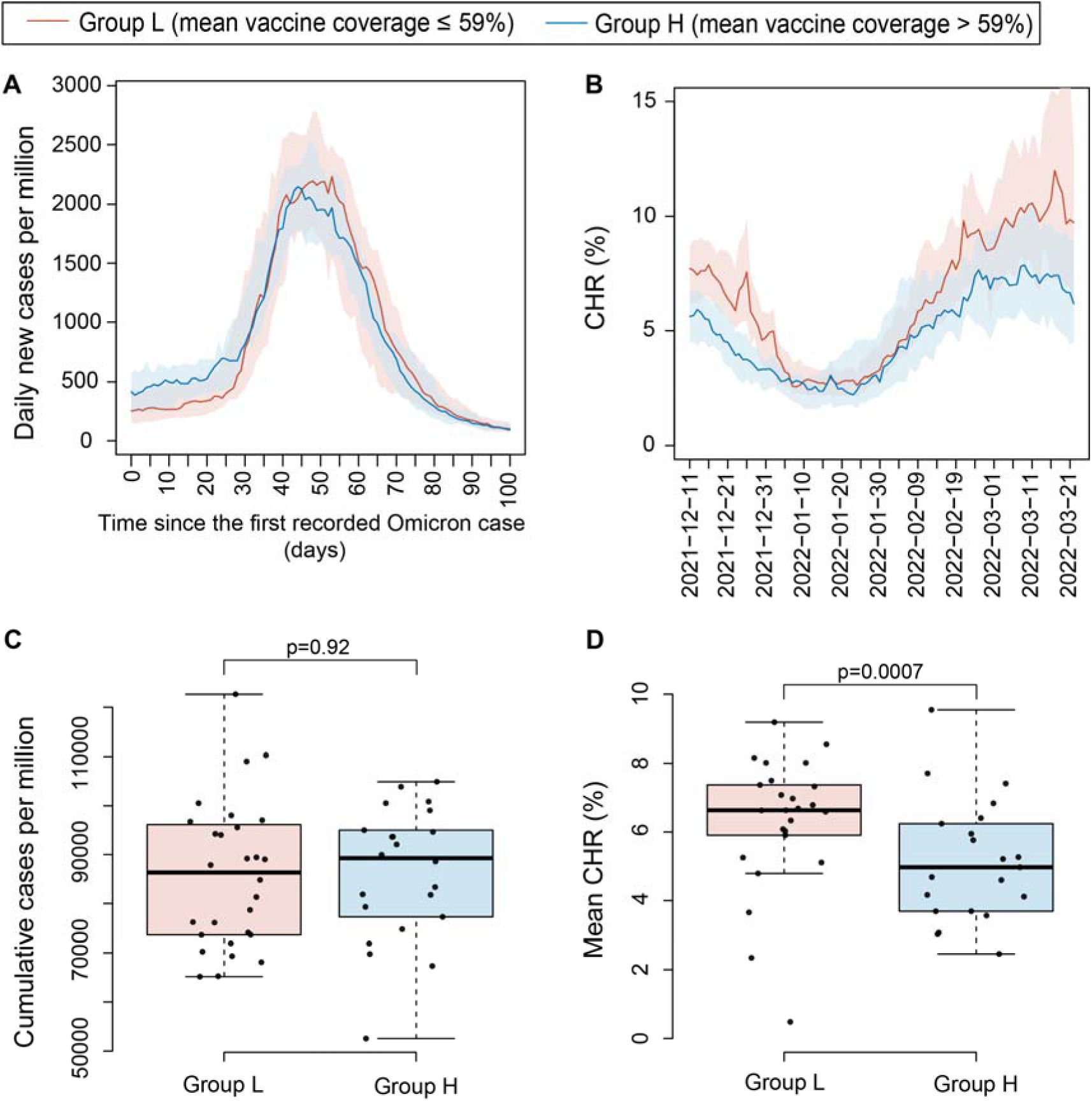
Comparisons of the COVID-19 transmission and CHR between the states with high and low vaccination coverages. (AB) The daily number of cases per million and daily CHR for the states with low vaccination coverage (Group L) and with high vaccination coverage (Group H). The red and blue lines represent the median value of Group L and H, respectively, and the shaded areas represent the interquartile range. (CD) Box plots of cumulative cases per million and average daily CHR during the first Omicron wave for states in Group L and Group H.

In contrast, in the Delta wave, Group H had significantly lower infection incidences than Group L (Figure S8), suggesting Omicron’s greater ability to evade immunity. While transportation and population mobility could also influence infection incidences, we observed that they were not the primary factors during the Omicron wave. Specifically, Group H had lower population mobility compared to Group L (Figure S7BD), and there was no significant difference in the timing of Omicron’s arrival between the two groups (Figure S9).

Moreover, we found that Group H had a significant lower average daily CHR compared to Group L (T-test, P-value<0.05, Figure 3BD), which highlighted the beneficial impact of vaccination on reducing hospitalizations. Interestingly, we observed a higher proportion of Omicron mutations in Group H compared to Group L during the growth of the outbreaks (as indicated by the grey shaded area in Figure 4A-C). When we applied a logit transformation to these proportions, similar patterns were observed (Figure S19). This can be explained by the transmission advantage of the Omicron mutations in the high-vaccine-coverage states. Hence, the lower CHR observed in Group H was likely to be attributed to both the protective effect of vaccination and the increased proportion of the Omicron variants.

**Figure 4.**
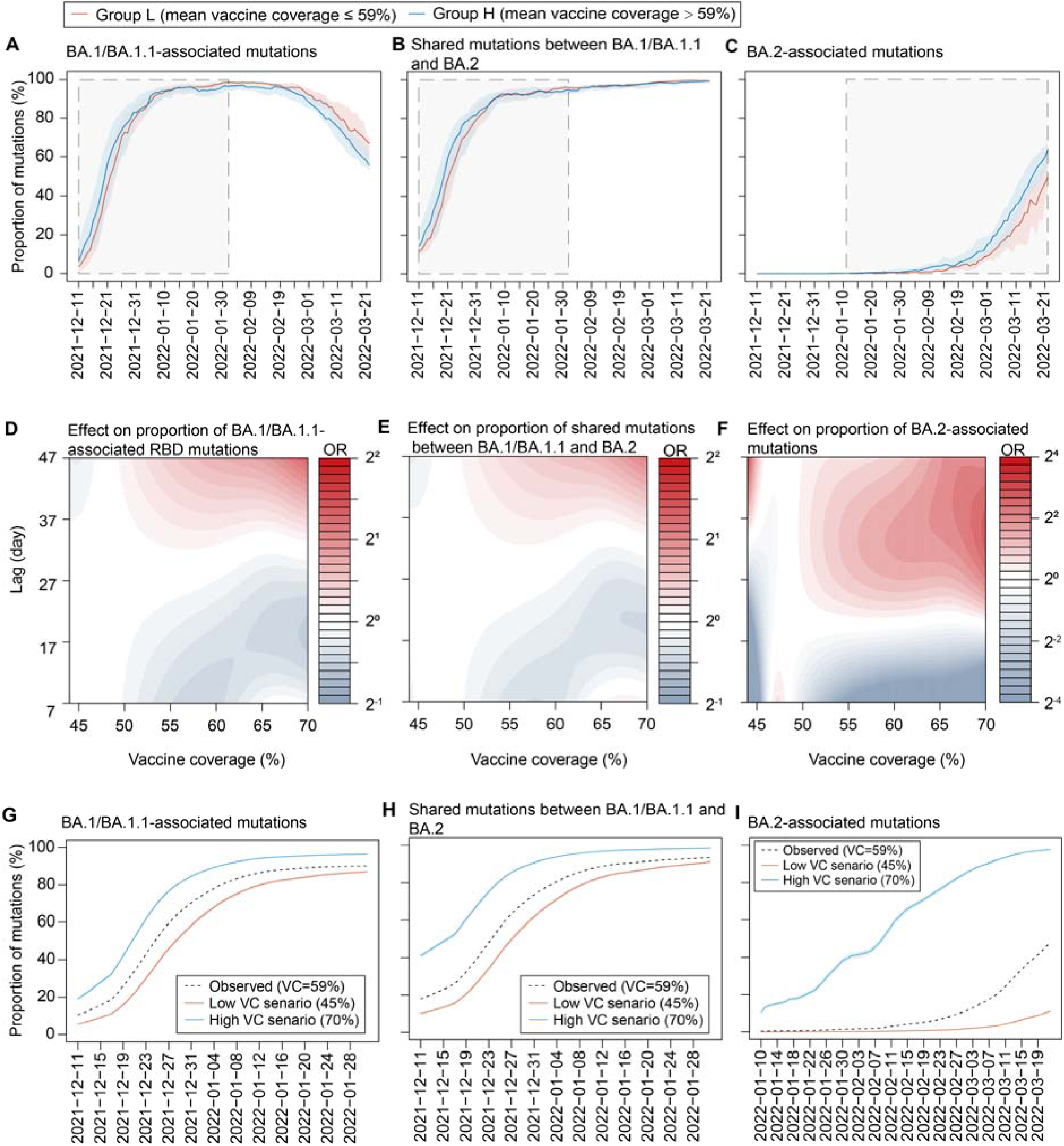
Estimated effects of vaccination coverage on the transmission of Omicron mutations from the intermediate models. (A-C) Mean proportion of BA.1/BA.1.1- associated mutations, shared mutations between BA.1/BA.1.1 and BA.2 subvariants, and BA.2-associated mutations in Group L (with low vaccination coverage) and Group H (with high vaccination coverage) states. The grey-shaded areas represented the period during which the proportion of the mutation grew. The mean proportion of BA.1 associated mutations is the average daily proportion of S371L, G496S, and G446S; mean proportion of BA.2- associated mutations is the average daily proportion of S371F, T376A, R408S, and D405N; mean proportion of shared mutations between BA.1/BA.1.1 and BA.2 is the average daily proportion of G339D, S373P, S375F, K417N, N440K, S477N, T478K, E484A, Q493R, Q498R, N501Y, and Y505H. (D-F) Relationships between vaccination coverage and the proportion of three types of mutations at different time lags. Redder colors indicated higher OR of mutation proportion. (G-I) The estimated proportions of BA.1/BA.1.1-associated mutations, shared mutations between BA.1/BA.1.1 and BA.2 subvariants, and BA.2- associated mutations under scenarios of constant vaccination coverage (VC) at 45% and 70%. The observed proportion of mutations (indicated by the dashed line) under the actual VC conditions (with an average value of 59%) was assumed as the reference point.

To further explore whether this transmission advantage could also be related to population mobility, we conducted an alternative study focusing on the period before vaccination (from January 23, 2020 to December 31, 2020). In this period, we detected a significant correlation between population mobility and the number of infections (Figures S10 and S11), suggesting that the initial transmission of COVID-19 was predominantly influenced by population mobility. Additionally, we found that both groups had low proportions of virus mutations (Figure S12), indicating that these mutations did not confer a clear transmission advantage in the absence of vaccination.

### Mediation effect of virus mutations on the vaccine-CHR relationship

We evaluated the mediation effect of virus mutations on the relationship between vaccination and the CHR using a mediation analysis with three specific models: *Basic (Total-effect)*, *Intermediate*, and *Mediation (Direct and indirect effects) models* (see Figure 1 and Methods).

After variable selection (Table S4), the best-fitting *Basic model* demonstrated that a higher vaccination coverage was significantly associated with a lower CHR. A 25% increase in the coverage resulted in a reduction of CHR with odd ratio (OR) of 0.37 (95% CI 0.36-0.38), representing the total effect of vaccination coverage (Table 1).

**Table 1.** Effects of factors on COVID-19 CHR in the *Basic model* and *Mediation model*. The *Basic model* estimated the total effect of vaccination on the CHR. The *Mediation model* estimated both the direct effect of vaccination and the mediation effect of the proportion of Omicron mutations on the CHR.

|  | *Basic model*<br>(CHR~vaccine)<br>**OR**<br>(95%CI) | **P value** | *Mediation model*<br>(CHR~vaccine+virus mutation)<br>**OR**<br>(95%CI) | **P value** |
| --- | --- | --- | --- | --- |
| **Vaccine** |  |  |  |  |
| Vaccination coverage (25%) | 0.369<br>(0.355, 0.383) | <0.001 | 0.853<br>(0.754, 0.966) | 0.012 |
| **Virus mutations** |  |  |  |  |
| Mean proportion of BA.1/BA.1.1-associated mutations (20%) | - | - | 0.870<br>(0.839, 0.901) | <0.001 |
| Mean proportion of BA.2-associated mutations (20%) | - | - | 0.868<br>(0.852, 0.884) | <0.001 |
| Mean proportion of shared mutations between BA.1/BA.1.1 and BA.2 (20%) | - | - | 0.994<br>(0.975, 1.014) | 0.573 |
| **Confounders** |  |  |  |  |
| Temperature (5°C) | 1.088<br>(1.083, 1.092) | <0.001 | 1.084<br>(1.077, 1.091) | <0.001 |
| Relative humidity (5%) | 1.018<br>(1.015, 1.020) | <0.001 | 1.013<br>(1.010, 1.015) | <0.001 |
| Hospital beds (1000) | 1.003<br>(1.002, 1.004) | <0.001 | 1.003<br>(1.001, 1.004) | <0.001 |
| Weekend (Yes vs No) | 1.001<br>(0.997, 1.005) | 0.592 | 1.002<br>(0.998, 1.007) | 0.293 |
| Holiday (Yes vs No) | 1.030<br>(1.021, 1.039) | <0.001 | 1.021<br>(1.011, 1.030) | <0.001 |

The *Intermediate model*, which measured the impact of vaccination on the logit of the frequency of a newly adapted variant (see Methods), found a significant positive correlation between vaccination coverage and the proportion of Omicron mutations with about one-month lag. For example, a 10% increase in vaccination coverage (from 50% to 60%) was associated with a higher proportion of Omicron mutations at a lag of 47 days. The corresponding ORs for the proportions of BA.1/BA.1.1 mutations, shared mutations between BA.1/BA.1.1 and BA.2, and BA.2 mutations were 1.45 (95% CI 1.44, 1.46), 1.33 (95% CI 1.32, 1.34), and 1.49 (95% CI 1.46, 1.52) (Figure 4D-F). A lag of approximately one month might be explained by those compensatory mutations are often required to restore or increase the virus’s fitness ^27,28,29^. A previous study observed compensatory epistasis in the Omicron BA.1 variant contributed to maintaining its affinity to the angiotensin-converting enzyme 2 (ACE2) receptor ^29^.

We projected the spreads of these mutations under two extreme scenarios: low coverage (45%) and high coverage (70%). We used the observed average coverage of 59% as a reference (Figure 4G-I). The high coverage scenario showed a 20-30% higher in the proportion of BA.1/BA.1.1-associated mutations or shared mutations between BA.1/BA.1.1 and BA.2 subvariant during their growth, compared with the low coverage scenario.

Furthermore, the proportion of BA.2-associated mutations rose over twice as quickly in the high coverage scenario than the low coverage.

The *Mediation model* found that a 25% increase (from 45% to 70%) in vaccination coverage was directly correlated with a lower CHR, with an OR of 0.85 (95% CI: 0.75, 0.97) (Table 1). Indirectly, the model found this increase in the coverage corresponded to a 20% increase in the proportion of BA.1/BA.1.1-associated mutations (Figure 4G), which were associated with a lower CHR with an OR of 0.87 (95% CI:0.84, 0.90) (Table 1). Together, the rise in vaccination reduced CHR from 5.8% to 4.4% (see details in Supplementary Material), and even excluding direct vaccine protection, CHR still decreased from 5.8% to 5.1% due to the adaptation of Omicron variant as marginal effect. Similar patterns were observed with the shared mutations between BA.1/BA.1.1 and BA.2 subvariants (Figure 4H), and BA.2- associated mutations (Figure 4I and Table 1). Overall, we found that all relationships in the three models (represented by the paths in Figure 1) were statistically significant, and the direct effect of increased vaccination on CHR was smaller than the total effect. Meeting the criteria for classical causal mediation (see Methods) ^22^, the results suggested that the adaptation of Omicron mutations acted as a mediator in the relationship between vaccination coverage and CHR. We also conducted an alternative approach using Structural Equation Modeling to validate our model and findings, showing consistent results (see Supplementary Material).

Therefore, a higher vaccination coverage was associated with a higher proportion of immune-escape variants. When these variants are less virulent, resulting in less severe infections compared to previous variants or wild types, they can contribute to a lower CHR (Figure 5). Conversely, the emergence of more virulent strains may potentially counteract the effect of vaccine protection (see Figure S20 for a hypothetical scenario).

**Figure 5.**
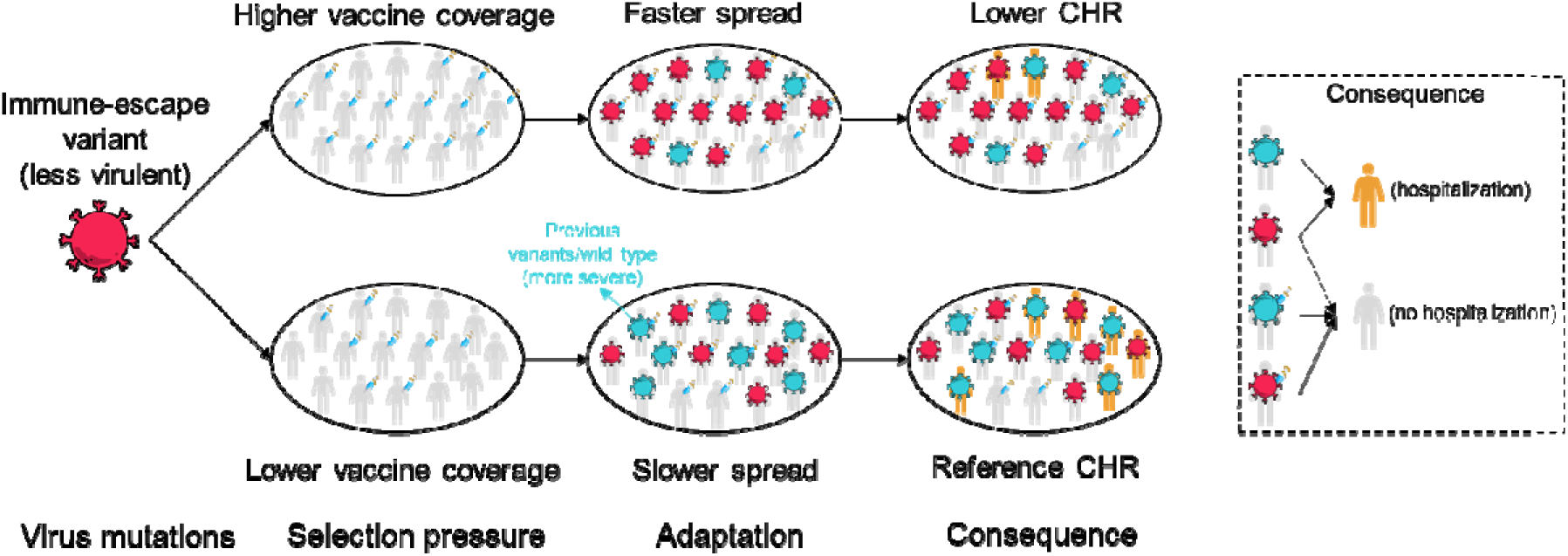
**The impact of vaccination on enhancing transmission potential of new immune-escape variant and the resulting impact on the overall CHR**. This figure illustrated the impact of vaccination on the adaptation of emerging immune-escape variants and its consequential effects on the CHR. High vaccination coverage might create a selection pressure that accelerate the rapid spread and adaptation of new immune-escape variants. If these variants are less virulent than the wild type or previously observed variants, a corresponding reduction in the CHR is anticipated.

### Effects of confounders

In addition to vaccination coverage and viral mutations, our analysis revealed additional factors that had an impact on CHR. We observed negative associations between CHR and daily mean temperature, relative humidity, and the number of staffed beds in hospitals (Table 1 and Supplementary Material). Interestingly, during the course of the outbreak, we noticed a temporal pattern: a surge in the number of daily reported cases was linked to an immediate reduction in CHR, followed by a subsequent increase in CHR approximately ten days later (Figure S13B). We attribute this finding to the limited hospital capacity, which may lead to a delay in the admission of certain severe cases.

### Model validation and sensitivity analysis

We conducted an alternative approach using structural equation model to validate our model and findings. The results confirmed a statistically significant indirect effect of the vaccine on reducing CHR through increasing the proportion of Omicron mutations (see Figure S14 and Supplementary Material).

In the sensitivity analyses, we first examined the impact of decreased vaccine effectiveness against hospitalization, assuming the protection dropped to 80% of the original level six months after full vaccination (Table S5, Figure S15). Second, we incorporated the effect of pre-existing immunity into our analysis (Table S6, Figure S16). The results of these sensitivity analyses were consistent with our primary findings (Table 1 and Figure 4).

## Discussion

Our study found that higher vaccination coverage was associated with faster adaptation of the new VOC, subsequently affecting the overall CHR. After linking the evolution theory to a lagged regression model, the findings were derived from mediation analyses with mixed-effect models along with genomic sequences among 50 US states and the District of Columbia during the first Omicron wave. These results provide additional insights into healthcare preparation for future outbreaks.

Vaccination has been expected to influence variant replacement by natural selection, but to what extent remains unknown. Our model results showed that the increased vaccine-induced immunity (from 45 to 70% coverage) was likely to speed up the replacement of Omicron variants from few weeks (for BA.1 or BA.1.1) to few months (for BA.2) (Figure 4G-I).

Because the new Omicron variants were generally associated with less severe disease outcomes ^15^, CHR in the high-vaccine-coverage states was further reduced to a level similar to the direct protection from vaccines. Meanwhile, many Western and Central European countries (including the United Kingdom) had about 10% higher vaccination coverage than the US by late 2021 ^30^. The earlier spread of Omicron, especially BA.2, has been observed in many of these countries than the US. Vaccines, were likely to play an important role, at least in part, in this early spread of BA.2.

Understanding the effect of vaccine on the VOC adaptation is important in decision-making in healthcare preparation and vaccine strategy ^4^. Our results raised an important question: What is the role of vaccines in balancing the prevention between massive transmission and severe outcomes? Designing vaccine strategies should consider not only the effectiveness and waning protection of vaccines but also the impacts on the spread of newly emerging variants. The study offers new insight into vaccine strategy for reducing hospitalization risk by shortening [or maintaining] the circulation of more [or less] virulent variants.

Cautionary insights arise from the study, highlighting that while vaccines provide direct protection against CHR, their indirect effect via the immune selection may sometimes be less beneficial or even result in negative impacts in certain scenarios. For example, in cases where a new VOC is more severe than its predecessor, a faster spread of this VOC selected by high partial population immunity can potentially increase CHR (Figure S20). Therefore, vigilant surveillance of the severity of new strains becomes critical to assess the healthcare system’s demand, even in regions with high vaccination coverage.

Our results suggested that vaccine effectiveness studies could consider the dynamic interplay between vaccination coverage rate and viral evolution. In addition to the waning immunity, the change in vaccine effectiveness might be affected by the emergence of new variants ^31^.

Adjusting for the heterogenicity of vaccination coverage (such as county-level vaccination rate) seems to be important to measure the waning of immunity as the model used by Lin et al ^31^. Similarly, vaccine strategy modelling studies may incorporate the mechanism of the replacement of circulating viruses to predict its impact on healthcare system.

This study has several limitations to consider. First, certain confounders, such as air pollution, personal self-medical treatments and under reporting might also impact CHR. Second, while our study did not directly incorporate infected travelers between countries and states, we observed no significant differences in the arrival times of Omicron between states (Figure S9). Our model incorporated the number of cases as a confounder, representing the combined effect of traveling and social distancing. Third, it is important to note that our focus was restricted to evaluating the impact of vaccination coverage on the adaptation (replacement) of new mutations. We did not explore the rate of the emergence of viral immune escape variants under vaccination pressure.

## Data Availability

All data produced in the present study are available upon reasonable request to the authors

## Acknowledgement

We thank City University of Hong Kong and Hong Kong Institute for Advanced for supporting this work, and we thank all of the researchers for generating and sharing genome data openly via GISAID. We thank Prof. Sarah Cobey at the University of Chicago for her valuable comments and contributions, which greatly enhanced the quality of our research.

## Authors contributions

Jingbo Liang collected epidemiological data, conducted the analyses, and wrote the first draft of the manuscript. Zhaojun Ding collected the genomic sequential data, calculated the mutation prevalence. Qingpeng Zhang provided important comments, and edited the manuscript. Hsiang-Yu Yuan conceptualized and designed the study, provided important comments, and edited the manuscript. All authors read and approved the manuscript.

## Ethics approval

Not applicable. No formal ethical review and informed consent were required, as the data used in this analysis were collected from online open datasets and all data were de-identified and aggregated to the state-level as described in the manuscript.

