## Supplementary file for "Impacts of vaccination on the variant selection of SARS-CoV-2 and the subsequent case hospitalization rate: insights from the first U.S. Omicron wave"

#### 1. Supplementary Method

##### 1.1 SARS-CoV-2 genomic surveillance data management

To ensure the quality of multiple sequence alignment, certain sequences were excluded, such as (i) those with more than 5% ambiguous amino acids and (ii) those with sequence lengths shorter than 1263 and longer than 1273. After these exclusions, 2,383,131 sequences were left for analysis. Multiple sequence alignments were performed using the Clustal Omega tool [1], with the reference sequence of the SARS-CoV-2 Wuhan-Hu-1 isolate (GISAID accession ID name called '*China/Wuhan-Hu-1/2019|EPI_ISL_402125*').

Using the genomic sequence data, we calculated the daily proportion of specific Omicron mutations within each state. This was assessed as the fraction of genomic sequences featuring that targeted mutation among all sequences submitted from the same state each day. Our analysis focused on 19 key mutations on the Receptor Binding Domain (RBD) of the Omicron variant[2], which were grouped into three categories: (i) BA.1/BA.1.1-associated mutations, including S371L, G496S, and G446S, (ii) shared mutations between BA.1/BA.1.1 and BA.2, including G339D, S373P, S375F, K417N, N440K, S477N, T478K, E484A, Q493R, Q498R, N501Y, and Y505H, and (iii) BA.2-associated mutations, including S371F, T376A, R408S, and D405N (Figure 2D and Figure S5).

##### 1.2 Adjusted effective vaccination coverage considering boosting and waning immunities

We calculated the effective vaccination coverage ($Vac_i(\mathrm{t})$) by summing the coverage after adjusting for vaccine waning and booster effects [3]. The calculation was performed as follows:

$$Vac_i(\mathrm{t}) = E_1 Vc_i(\mathrm{t}) + E_2 Vb_i(\mathrm{t}) + E_3 Vw_i(t). \qquad \text{(eq.1)}$$

In the i$^{th}$ state (i = 1, 2, 3, ⋯, 51) on the t$^{th}$ (from 11 December 2021 to 22 March 2022) day, the effective vaccination coverage ($Vc_{i,t}$) was calculated as the sum of three components: $E_1Vc_i(\mathrm{t})$, $E_2Vb_i(t)$, and $E_3Vw_i(t)$. $Vc_{i,t}$ represents the proportion of people fully vaccinated within six months; $Vb_i(\mathrm{t})$ represents the proportion of individuals fully vaccinated over six months who had received a booster dose (assuming that only those fully vaccinated over six months were eligible for a booster dose); and $Vw_i(t)$ represents the proportion of people fully vaccinated over six months who had not received a booster. The effectiveness ($E_1$, $E_2$, and $E_3$) against hospitalization for these groups was assumed to be 95%, 95%, and 91%, respectively [4].

##### 1.3 Case hospitalization rate (CHR) as a critical outcome metric

In this study, the case hospitalization rate (CHR) was used as a primary outcome metric, owing to its significant role in assessing hospitalization numbers and healthcare resource requirements. Infectious disease models are instrumental in forecasting the dynamics of case incidence. To get the number of hospitalizations, a key variable "proportion of cases that develop more severe disease requiring hospitalization" is needed in modelling (e.g. this variable was used by various researchers, including Imai et al.[5]). However, this proportion is often difficult to estimate. To address this challenge, CHR was used to represent this proportion.

CHR has been used to represent disease severity in many previous studies. For example, in an influenza study in the US [6], different CHR values were used to represent scenarios of low and high severity during influenza outbreak. Our study assumed that CHR is largely influenced by the proportion of cases that develop more severe illness requiring hospitalization (like most of the previous public health studies) and the balance between demand (disease incidence) and capacity (number of hospital beds). From the model projection perspective, this can be used as a metric to assess both disease severity and healthcare demand.

We investigated how the CHR changed during the first Omicron wave by calculating the average daily CHR before and after the peak (designated as $CHR_{before}$ and $CHR_{after}$, respectively). Given different states reached their peak at different times, we calculated the daily CHR before the peak, starting from when the daily reported cases exceeded a threshold (i.e., 25% of the peak number of daily cases in that state), and ending at the peak. Similarly, we calculated the daily CHR after the peak, beginning at the peak time and ending when the daily reported cases decreased below the threshold. To quantify the change in the daily average CHR before and after the peak, we calculated the percentage change as $(CHR_{after} - CHR_{before})/CHR_{before}$.

##### 1.4 Determine the total effect of vaccination coverage on virus mutations (*Basic Models*)

In our research, we used state-level population data to perform causal mediation analysis [7–9]. This statistical method can decompose the "total effect" of an exposure into two distinct components: the "direct effect" and the "indirect effect" [10,11]. These terms and the methodology have been widely used in both observational study [9,12] and randomized controlled trials study [13]. Within the scope of our investigation, this analytic framework can help not only assess the protective effect of vaccines (as established in previous studies) but also quantify the potential indirect impact of vaccine on the spread of immune escape mutations, subsequently influencing the CHR.

We developed the *Basic model* to obtain the total effect of vaccination on the CHR. We assumed that the risk of hospitalization for COVID-19 infections follows a binomial distribution with the rate of CHR. The variables used in the *Basic Model* are described in detail in the main text. We incorporated the log-transformed daily number of reported cases ($Log(cases)_{i,t}$) into the model based on its linear relationship with the CHR. To capture the lag-response relationship between the daily cases and CHR, we used a cross-basis function, $cb(Log(cases)_{i,t},$ lag), with lag periods ranging from a minimum of 0 days to a maximum of 14 days. To estimate the Odds Ratio (OR) of the CHR, we calculated the exponential transformation of the corresponding coefficients.

##### 1.5 Determine the effect of vaccination coverage on transmission of virus mutations (*Intermediate Models*)

In the intermediate model, we explored how vaccination impacts the selection coefficient for the Omicron variant. Here, we explained how evolutionary process was modelled by the equations for the changes in the variant ratio. According to the previous study [14], the selection coefficient, denoted by $s$, is defined as the rate of change in the logit-transformed frequency of a new variant over time: $s = \frac{dlog\left(\frac{p(t)}{1-p(t)}\right)}{dt}$, where $p(t)$ is the proportion of the new variant at time $(t)$, and $1 - p(t)$ is the proportion of the other pre-existing variants (see Equation 2.1 at [14]).

Selection coefficient s is set to be proportional to the percentage of the hosts who are vaccinated (i.e. effective vaccine coverage VC(t) as we used) [15]. Then we have

$$s = \frac{dlog\left(\frac{p(t)}{1-p(t)}\right)}{dt} = \beta \cdot VC(t) \quad \text{(eq.2)}$$

where $\beta$ represents the effect of vaccination on $s$. Integrating this over time yields:

$$log\left(\frac{p(t)}{1-p(t)}\right) = \int_{to}^{t} \beta \cdot VC(t))dt \quad \text{(eq.3)}$$

where $to$ and $t$ represent the period during the variant replacement. If we rewrite this in discrete time, in terms of lag effect on variant selection, we have

$$log\left(\frac{p(t)}{1-p(t)}\right) = \sum_{t0}^{t} \beta \cdot VC(\tau) = \sum_{lag=0}^{t-t0} \beta \cdot VC(t - lag) \quad \text{(eq.4)}$$

where $lag$ is the lagged time and $\beta$ was assumed to be constant over time. If we further consider that $\beta$ can vary across different lagged time, the right term can be formulated using a distributed and nonlinear crossbasis function:

$$log\left(\frac{p(t)}{1-p(t)}\right) = \sum_{lag=0}^{Max} cb(VC(t), lag) \quad \text{(eq.5)}$$

where $cb$ is the crossbasis function and $Max$ is the maximum considered lag time. We further adapt this model to incorporate regional variations using a mixed-effects model, where $i$ represents each region and $\alpha_i$ represent region-specific random effect:

$$log\left(\frac{p_i(t)}{1-p_i(t)}\right) = \alpha_i + \sum_{lag=0}^{Max} cb(VC_i(t), lag) \quad \text{(eq.6)}$$

Furthermore, considering the effects of both vaccination and cases, we developed *Intermediate Models* as follows:

$$\log\left(\frac{P_{BA1,i,t}}{1-P_{BA1,i,t}}\right) = \alpha + \sum_{lag=7}^{47} cb\left(Vac_{i,t}, \text{lag}\right) + \delta_1 Log(cases)_{i,t} \quad \text{(eq.7)}$$

$$\log\left(\frac{1-P_{BA12,i,t}}{P_{BA12,i,t}}\right) = \alpha + \sum_{lag=7}^{47} cb(Vac_{i,t}, \text{lag}) + \delta_2 Log(cases)_{i,t} \quad \text{(eq.8)}$$

$$\log\left(\frac{1-P_{BA2,i,t}}{P_{BA2,i,t}}\right) = \alpha + \sum_{lag=7}^{47} cb(Vac_{i,t}, \text{lag}) + \delta_3 Log(cases)_{i,t} \quad \text{(eq.9)}$$

where $P_{BA1,i,t}$, $P_{BA12,i,t}$, and $P_{BA2,i,t}$ are the daily proportion of BA.1/BA.1.1-associated mutations, shared mutation of BA.1/BA.1.1 and BA.2 subvariants, and BA.2-associated mutations, respectively. In these *Intermediate Models*, the dependent variables, namely $\log\left(\frac{P_{BA1,i,t}}{1-P_{BA1,i,t}}\right)$, $\log\left(\frac{1-P_{BA12,i,t}}{P_{BA12,i,t}}\right)$, and $\log\left(\frac{1-P_{BA2,i,t}}{P_{BA2,i,t}}\right)$, are the logit transformations of the mutation proportions. The same logit term has been used to represent the selection coefficient on the vaccine escape mutations in a prior modelling study published in PNAS (see equation 1) [15]. We hypothesize that the vaccines' indirect effects on the CHR via the adaptation of immune-escape strains are driven by natural selection mechanisms. Moreover, $cases_{i,t}$ was also included to explore the possible influence on the selection coefficient of the Omicron immune escape mutations. To quantify these selection effects, we estimated the odds ratios (ORs) for the proportions of these three mutation types by exponentiating the corresponding coefficients derived from our models.

We also did an alternative Intermediate model using the cumulative cases instead of daily cases as confounders as follows: $\log\left(\frac{P_{Mut,i,t}}{1-P_{Mut,i,t}}\right) = \alpha + \sum_{lag=7}^{47} cb\left(Vac_{i,t}, \mathrm{lag}\right) + \delta_1 Cum\ cases_{i,t} + \delta_4 Cum\ cases_{i,t} \times Vac_{i,t}$ (eq.10). We found the alternative model results (Figure S17) were consistent with the main results (Figure 4).

**1.6 Control of confounding factors**

In our study, we adjusted for various confounding factors. The selection of confounders and independent variables for our study was based on two pivotal criteria: substantial evidence indicating their potential to influence disease severity and hospitalization rates, and the availability of reliable data.

First, we used mixed-effect models to account for state-level local effects, addressing variations in health status and healthcare access across different states. Second, recognizing the impact of non-pharmaceutical interventions like social distancing and mask-wearing on disease transmission, we indirectly accounted for their effects by incorporating the daily case numbers. Our analysis found a linear relationship between the logarithm of case numbers and CHR (Figure S13A). Third, we included hospital capacity data, as it indicates a hospital's patient admission capacity. Overwhelmed health systems may lead to outpatient management of cases that would otherwise require hospitalization, thus affecting CHR. Fourth, we included data of mean temperature and humidity, given their established association with disease severity. Fifth, we incorporated index variables for weekends and public holidays to adjust for temporal variations, accounting for differences in hospitalization behavior observed during these periods. Finally, we acknowledged the potential impact of public awareness on case reporting, but lacked specific U.S.-wide data. To address this, we assumed stable awareness levels within individual states and adjusted for inter-state reporting rate variations using a mixed-effects model. This approach also accounted for the possibility of underreporting, a common issue even in randomized control trials (RCTs) like the study by Lin et al [16].

##### 1.7 Alternative structural equation model

To validate the mediation effect observed in the main result, we employed an alternative structural equation model (SEM). This model was instrumental in quantifying the indirect influence of vaccination coverage. Specifically, we quantified the indirect effect of vaccination coverage by computing the product of the regression coefficients connecting the independent variable (vaccination coverage) to the mediator variable (virus mutations), and the mediator variable to the dependent variable (CHR). The model estimated total effect was subsequently calculated as a combined sum of the direct and indirect effects. We utilized the "*sem*" [17] package in R to construct this alternate model, thereby providing further validation to our results.

#### 2. Supplementary Result

##### 2.1 CHR projections for low and high vaccination coverage scenarios

We projected CHR under low (45%) and high (70%) vaccination coverage scenarios using results from our mediation analysis (Figure 4). We set a reference with an average vaccination coverage of 59%, which corresponded to a CHR of 5%. A decrease in vaccination coverage from 59% to 45% led to an increase in CHR, both directly (OR=1.093) and indirectly (OR=1.072), resulting in a CHR of 5.8%. Increasing the vaccination coverage from 45% to 70% was correlated with a decline in CHR both directly (OR=0.85) and indirectly (OR=0.870), leading to a CHR of 4.4%. Notably, the marginal effect of increasing vaccination coverage from 45% to 70% resulting in CHR reduced indirectly (OR=0.87) from 5.8% to 5.1%.

**2.2 Effects of other confounders on CHR**

During the Omicron wave, we observed an average daily mean temperature of 1°C and relative humidity of 62%, with interquartile ranges of -3°C-7°C and 58%-73%, respectively (Table S2). Our analysis indicates that an increase of 5°C in temperature is associated with a higher CHR (OR=1.08, Table 1), suggesting that the rise in temperature may have contributed to the increase in CHR after the peak. We also found that an increase of 5% in relative humidity is associated with a higher CHR (OR=1.01, Table 1). Furthermore, we observed a slightly higher CHR during public holidays than on workdays, with an OR of 1.021 (Table 1).

To investigate the lag effect of the number of daily cases on CHR, we found that a higher number of daily cases was associated with a lower CHR at short lags, but a higher CHR one week later. For instance, a large number of cases (i.e., log-transformed as 10) reduced the CHR compared to the reference number of cases (i.e., log-transformed as 6), with an OR of 0.21 (95% CI: 0.213, 0.225). This may be due to the hospital being overburdened with a large number of patients, causing some serious patients to not be admitted immediately. However, a large number of cases may result in a higher hospitalization rate one week later. For example, a large number of cases (i.e., log-transformed as 10) was associated with a higher hospitalization rate (OR of 1.42 (95% CI: 1.43, 1.46)) with a lag of 9 days compared to the reference number of cases. This is because some severe cases that could not be admitted immediately due to the limited hospital capacity may be admitted after a week's wait.

##### 2.3 Alternative structural equation model

We employed an alternative Structural Equation Model (SEM) to estimate and visualize the direct and indirect effects of vaccination coverage on the CHR, as presented in Figure S14. The arrows from V to M signify positive direct relationships between the independent variable V (vaccination coverage) and the mediator M (mutation proportion), as indicated by positive coefficients (e.g., 0.302 with a 95% CI of 0.161 to 0.443). Similarly, the arrows from M to H represent the negative relationship between the mediator M (mutation proportion) and the dependent variable H (CHR), as indicated by negative coefficients (e.g., -0.028 with a 95% CI of -0.030 to -0.026). Importantly, all these relationships were statistically significant with p-values less than 0.001. In summary, the alternative SEM aligns with our main findings, showing that higher vaccination coverage can indirectly reduce CHR by increasing the proportion of less severe Omicron variants.

##### 2.4 Changes in CHR before and after a peak

Figure S18B shows the percentage change in the average daily CHR from before to after the peak. We found that the CFR increased after the peak in both groups, but to a lesser extent in Group H than in Group L (T-test, P-value=0.04<0.05).

#### 3. Supplementary Figures

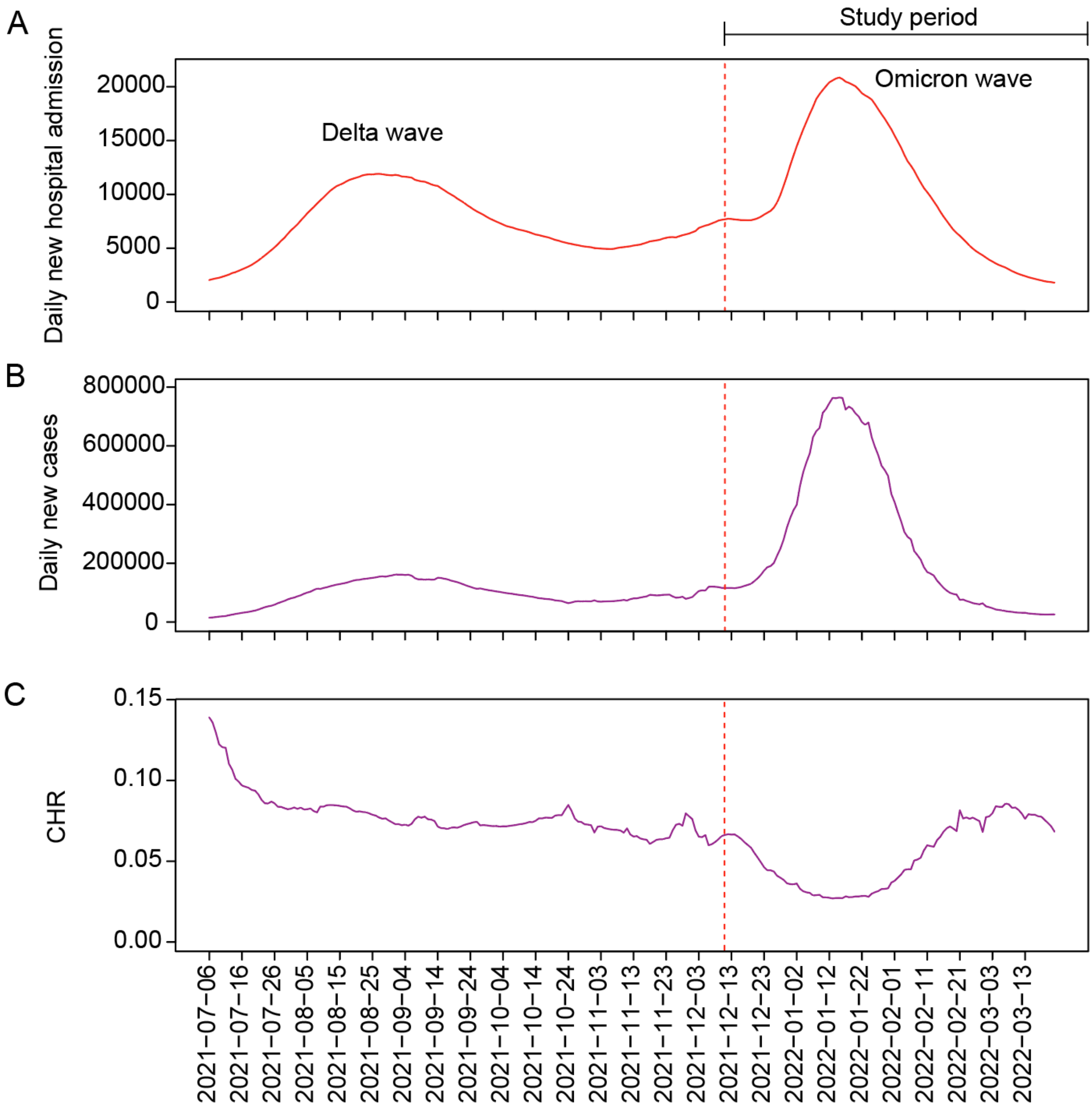


**Figure S1.** The number of daily hospital admissions, reported cases, and daily CHR during the Delta and first Omicron wave in the United States. The red line denotes the start time of the first Omicron wave when the proportion of the Omicron variant was over 5%.

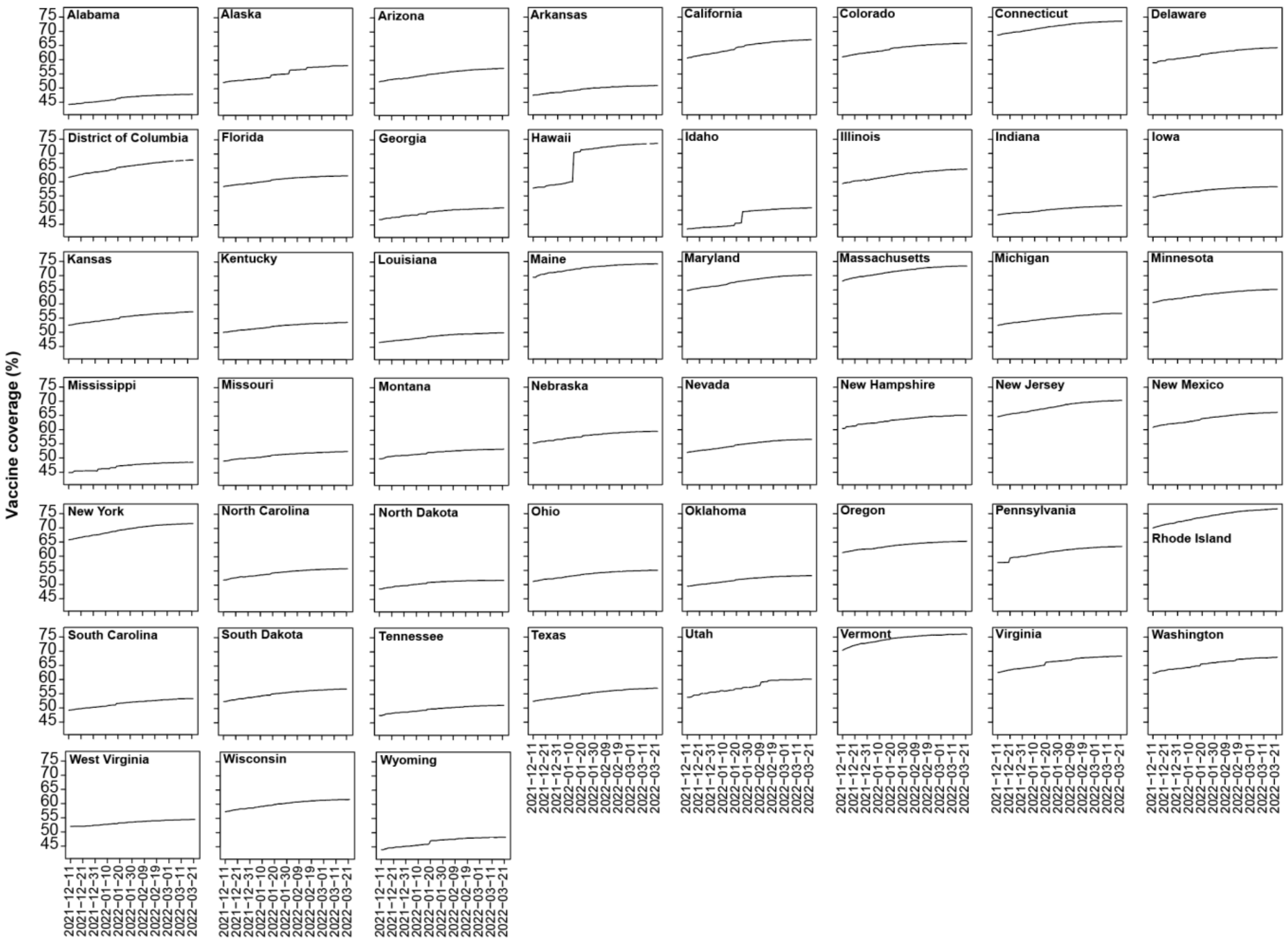


**Figure S2.** Time series of the cumulative proportion of people who were fully vaccinated at the state level during the first Omicron wave.

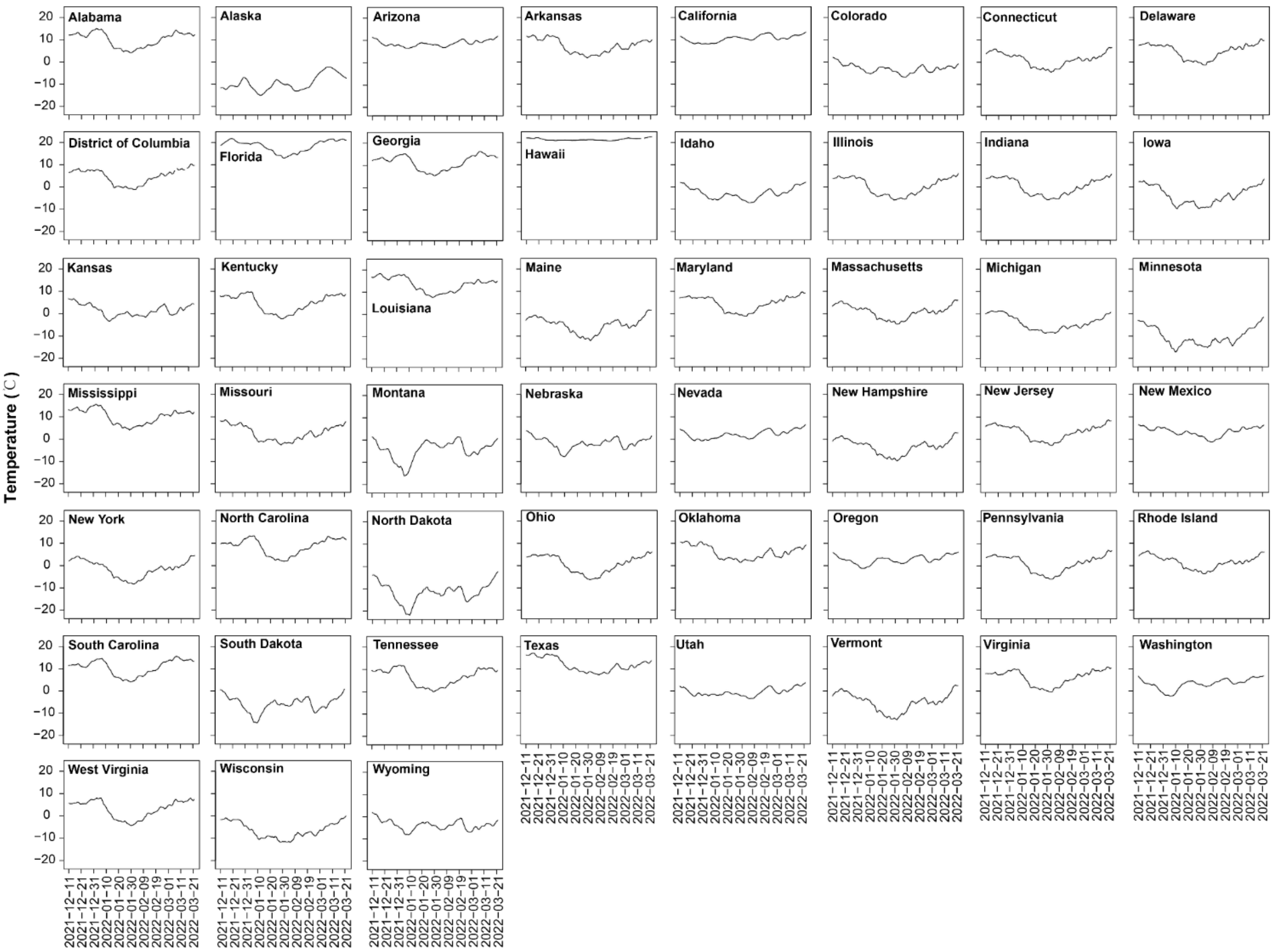


**Figure S3.** Time series of daily mean temperature at the state level during the first Omicron wave.

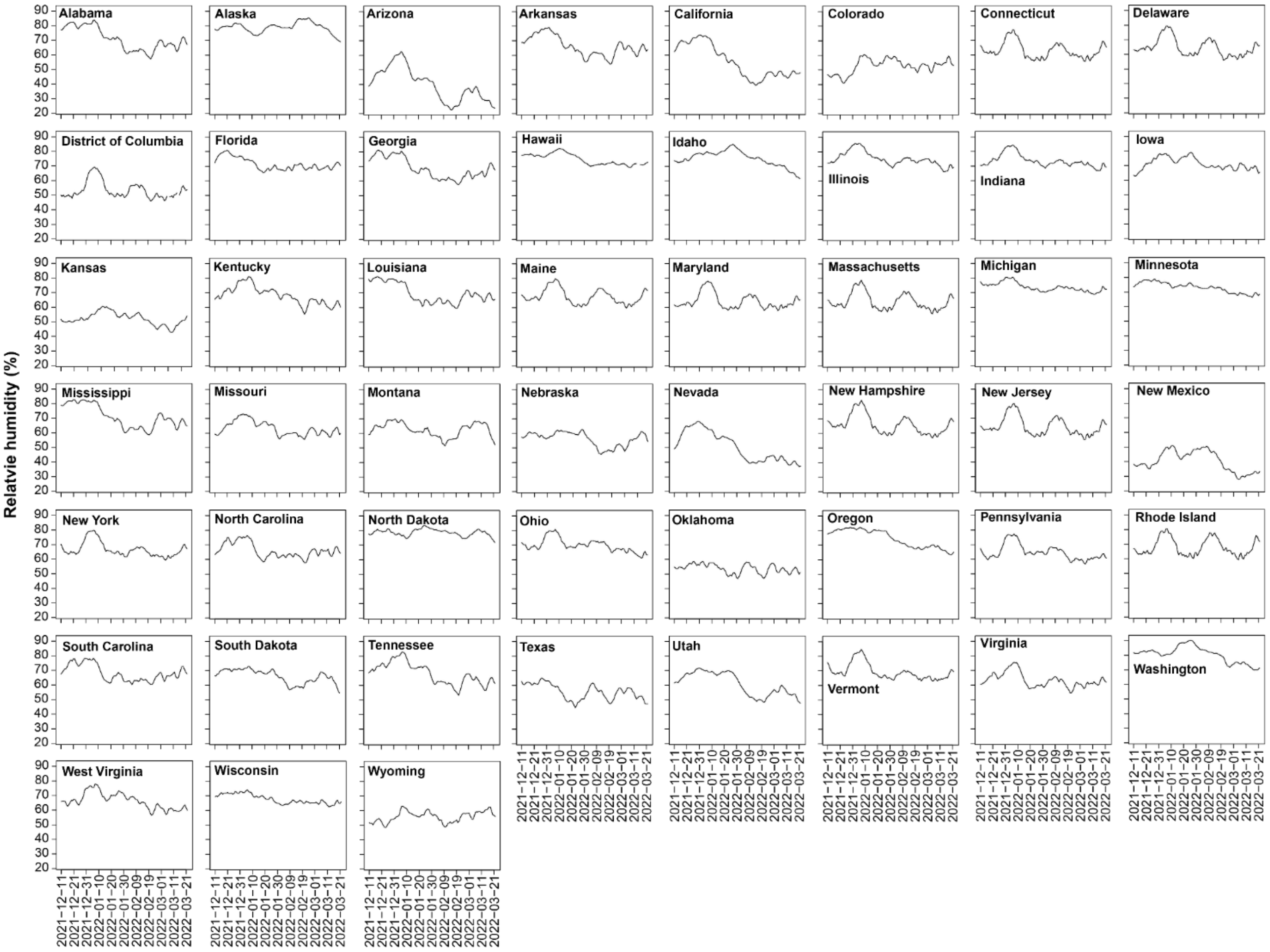


**Figure S4.** Time series of daily mean relative humidity at the state level during the first Omicron wave.

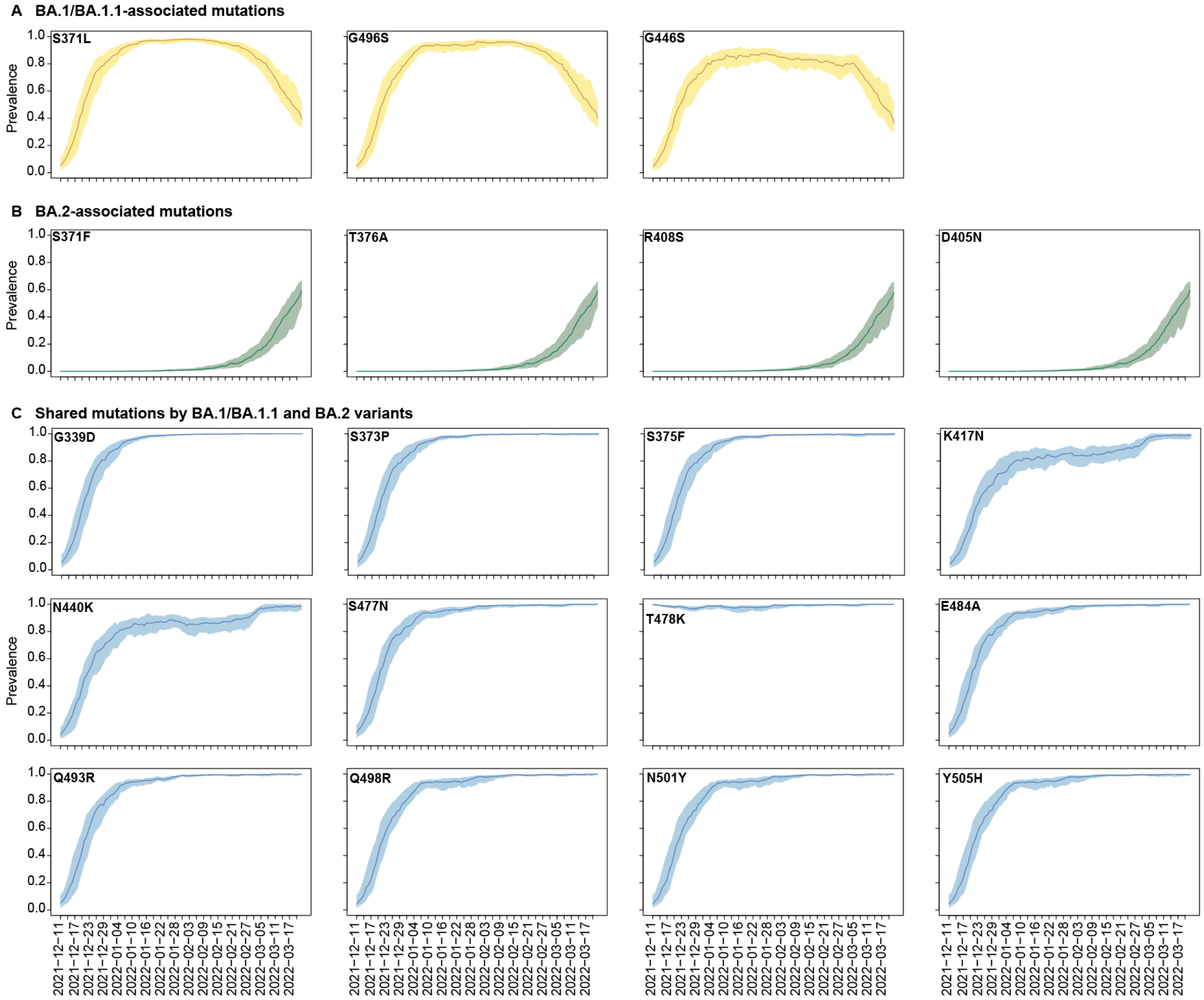


**Figure S5.** Prevalence of 19 RBD mutations of the Omicron variant in the United States during the first Omicron wave. The mutations are categorized as either associated with BA.1/BA.1.1 (A), BA.2 (B), or shared between BA.1/BA.1.1 and BA.2 variants (C). The dark-colored lines represent the median prevalence of each mutation, while the shaded areas represent the interquartile ranges.

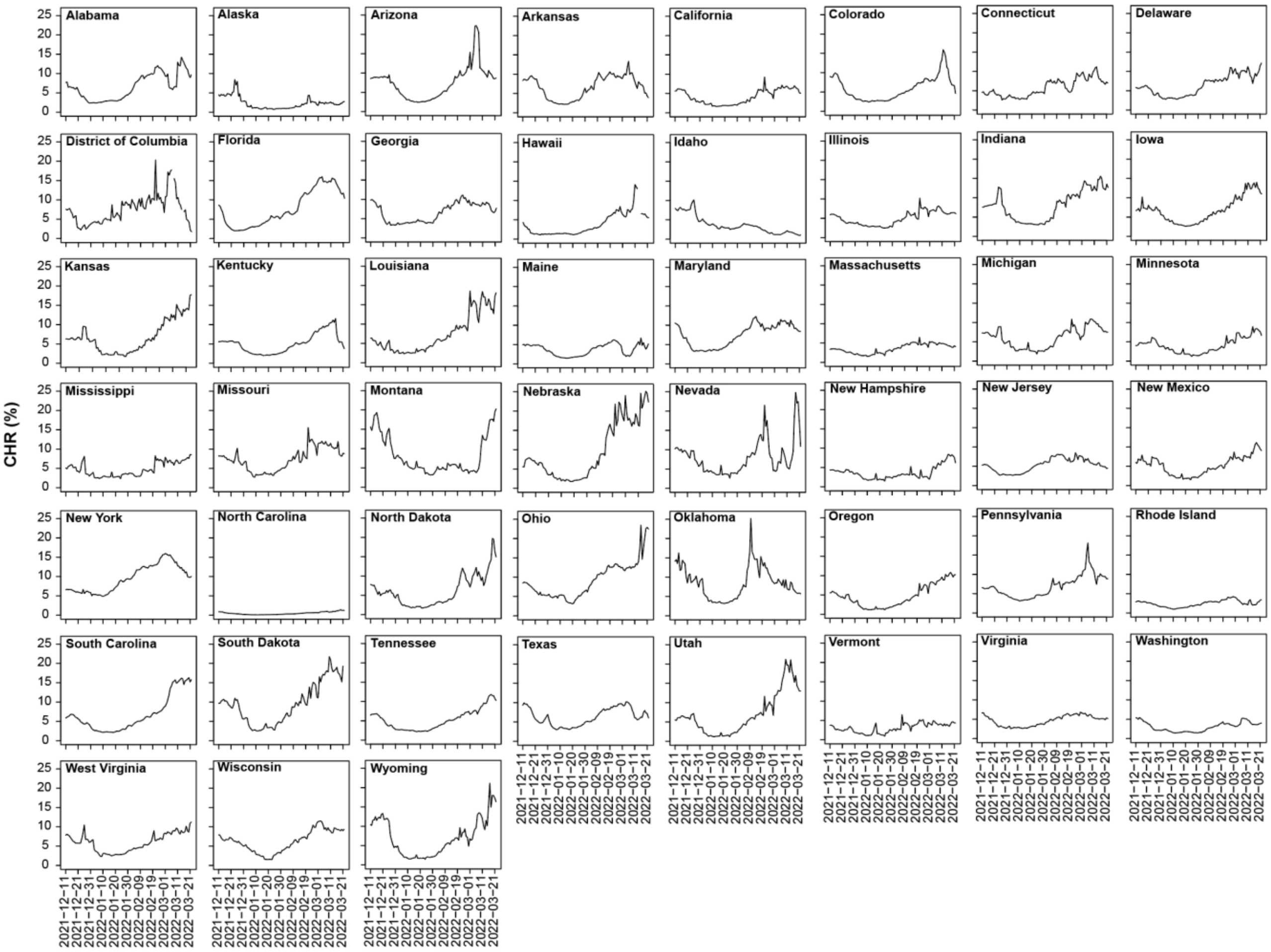


**Figure S6.** Time series of daily CHR at the state level during the first Omicron wave.

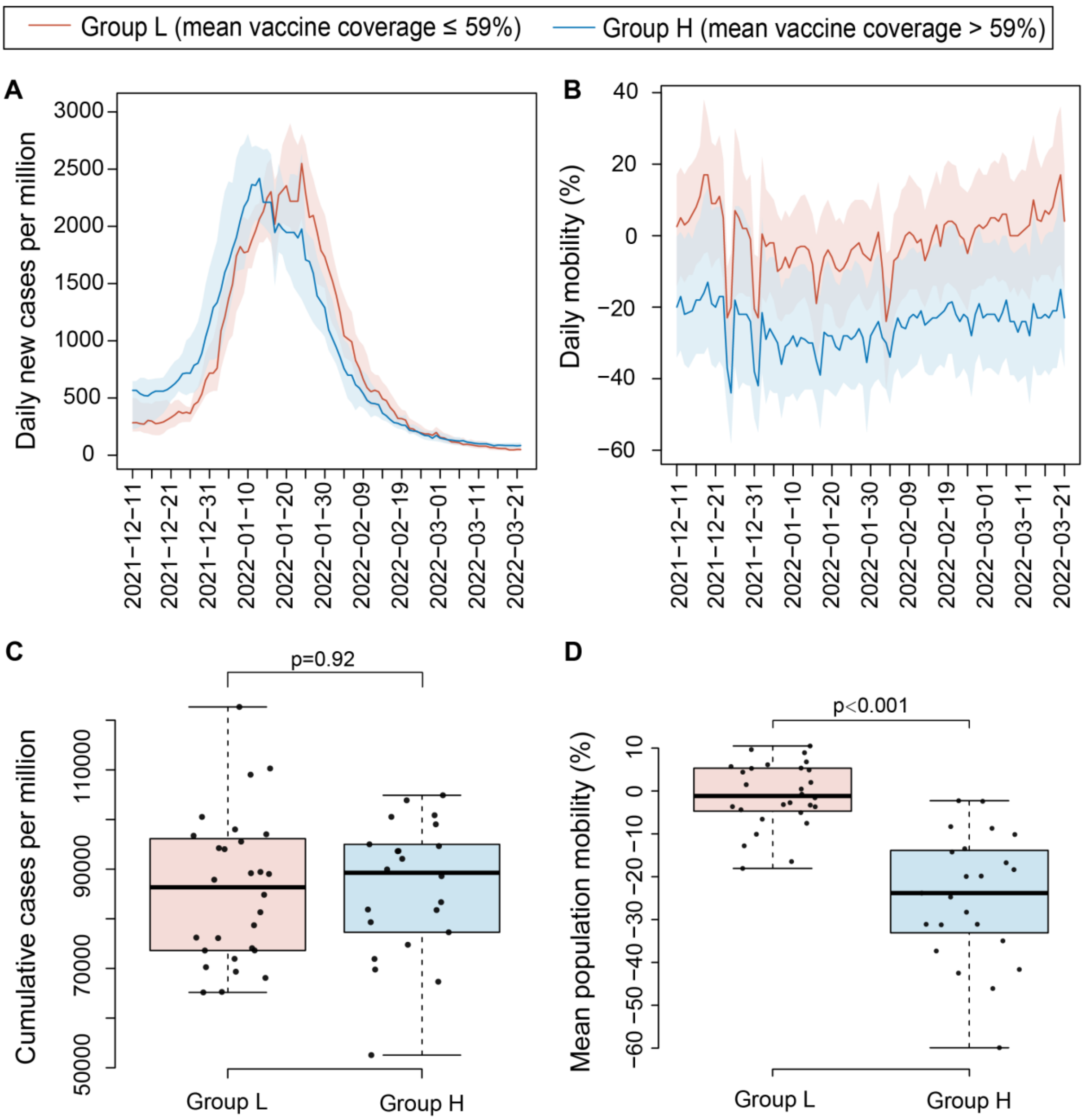


**Figure S7. Comparisons of the COVID-19 transmission and population mobility between the states with high and low vaccination coverages.** (AB) The daily number of cases per million and daily population mobility for the states with low vaccination coverage (Group L) and with high vaccination coverage (Group H). The red and blue lines represent the median value of Group L and H, respectively, and the shaded areas represent the interquartile range. (CD) Box plots of cumulative cases per million and daily average mobility during the first Omicron wave for states in Group L and Group H. The mobility represents the google transit stations percent change from the baseline (the baseline day is the pre-pandemic median value from the 5-week period between January 3 and February 6, 2020).

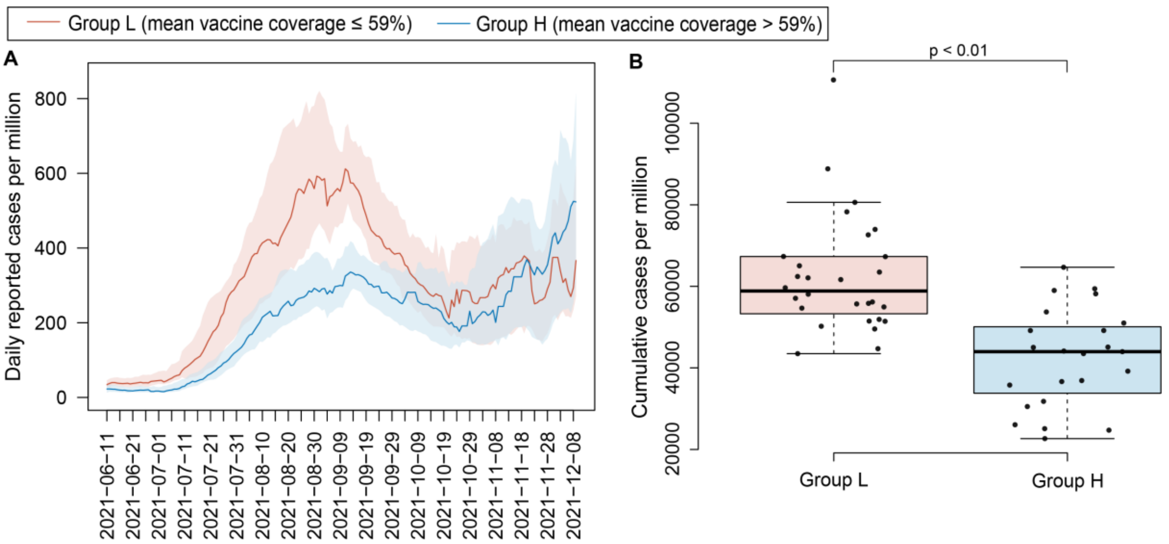


**Figure S8. Comparisons of infection incidence between the states with high and low vaccination coverages during the Delta wave.** (A) Daily number of cases per million population for states with low vaccination coverage (Group L) and with high vaccination coverage (Group H). (B) Box plot of cumulative cases per million during the Delta wave for states in the two groups.

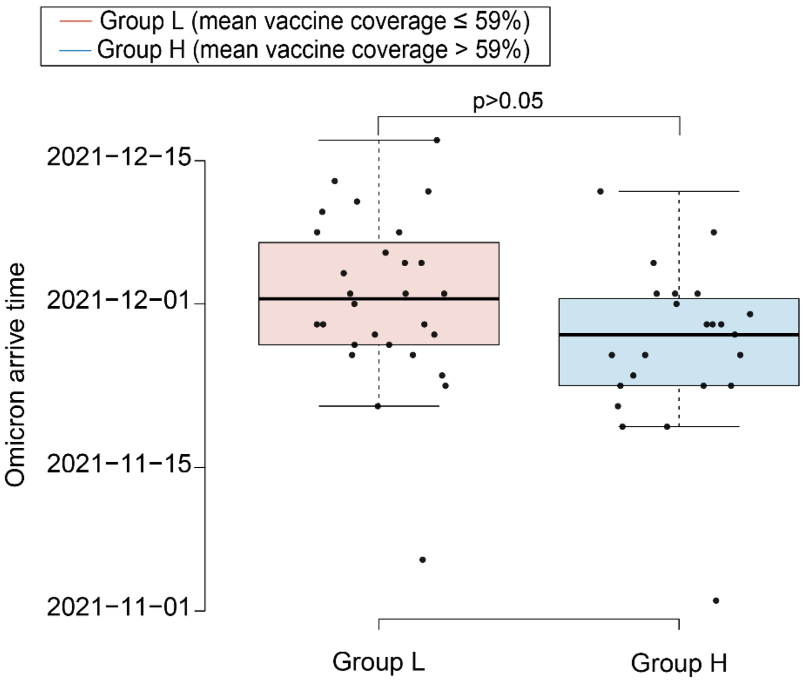


**Figure S9.** Arrival time of the Omicron variant in each state of the United States. The arrival time is defined as the earliest recorded day when the presence of the Omicron sequence was initially documented in the GISAID dataset for each respective state.

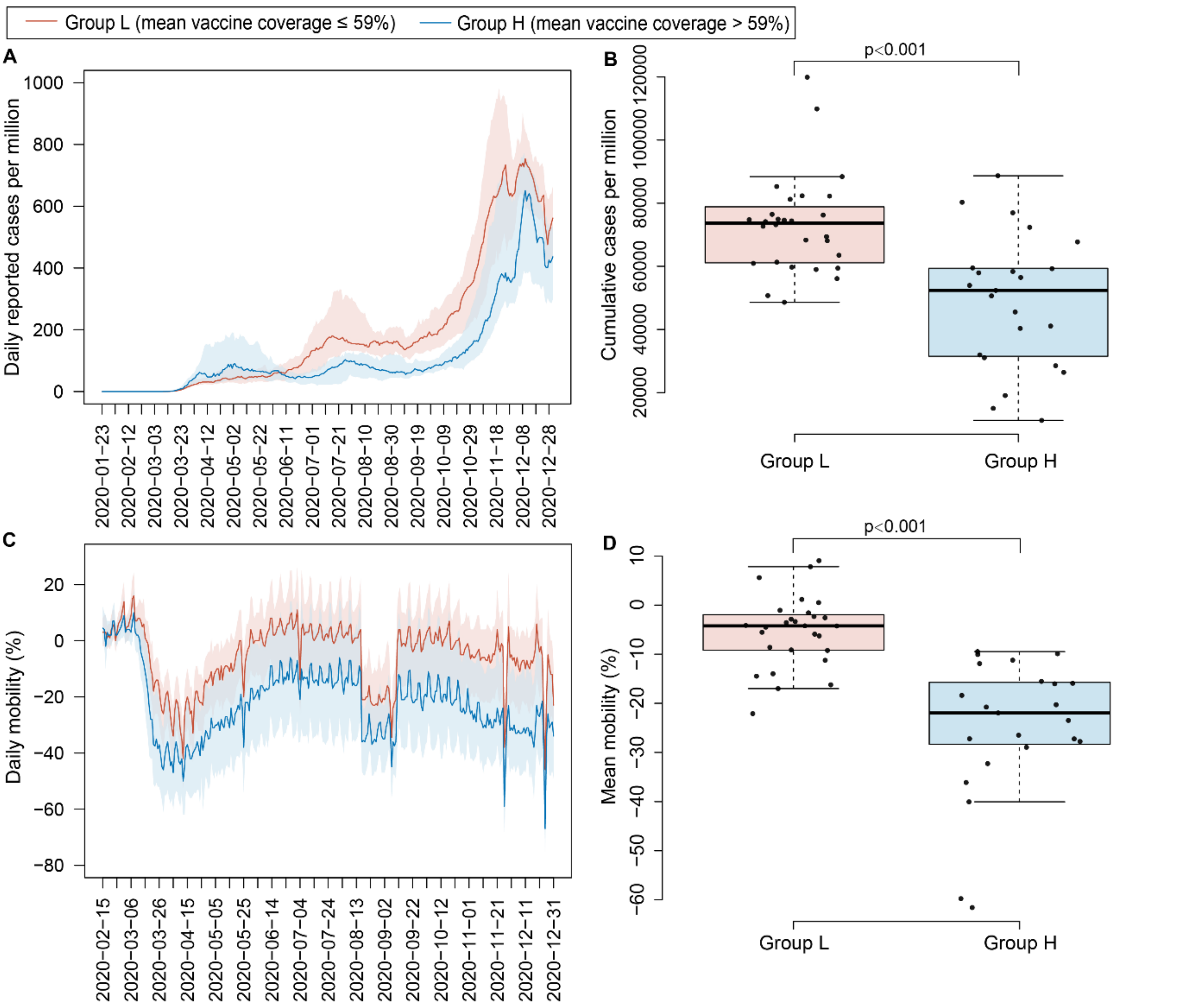


**Figure S10.** Comparisons of infection incidence and population mobility between the states with high and low vaccination coverages during the period when vaccination was not introduced (January 23, 2020 – December 31, 2020). (A) Daily number of cases per million population for states in Group L and Group H. (B) Box plot of cumulative cases per million during the period when vaccination was not introduced for states in the two groups. (C) Daily population mobility for states in Group L and Group H. (D) Box plot of daily average population mobility for states in the two groups.

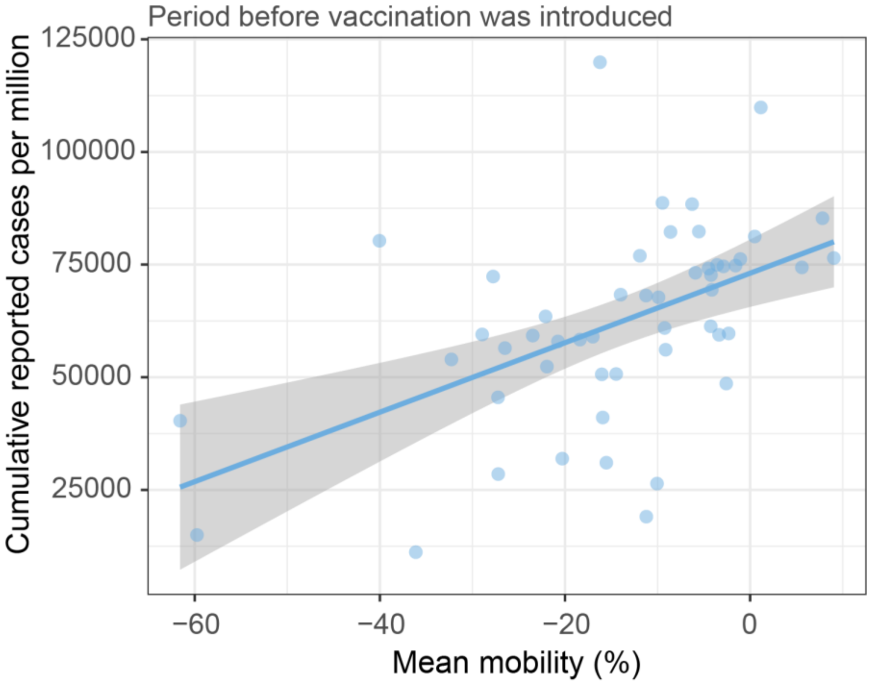


**Figure S11.** Scatter plot of the state-level daily average population mobility and the cumulative number of reported cases. Number of reported cases per million was shown between January 1, 2020 and December 31, 2020. The trend was depicted using a linear regression line, while the grey shaded region indicates the 95% confidence.

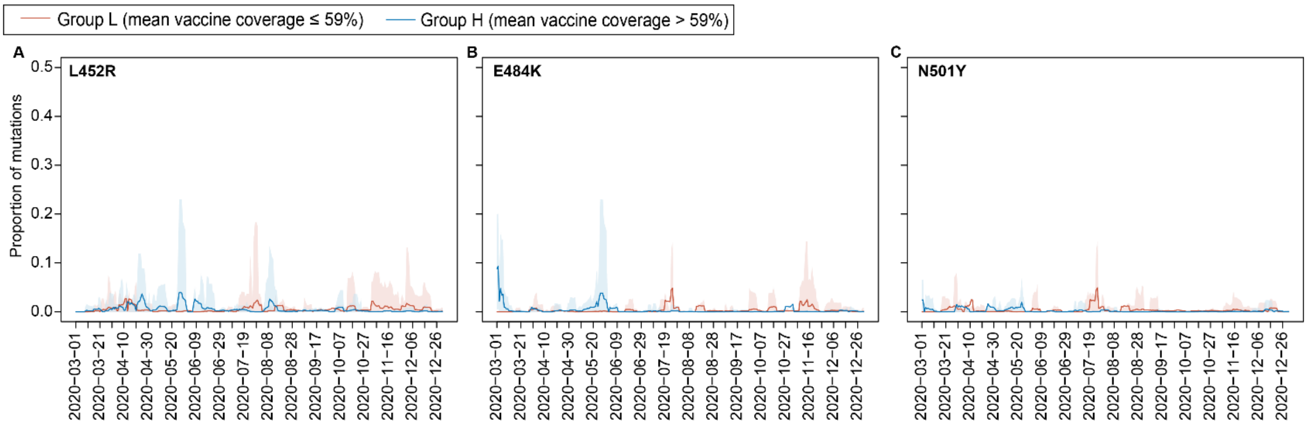


**Figure S12.** Comparison of the daily proportion of three key SARS-CoV-2 mutations. (A) L452R, (B) E484K, and (C) N501Y between states in two groups (Group L and Group H) during the period from January 1, 2020 to December 31, 2020, prior to the introduction of vaccines. The mean proportion for Group L is represented by the red line, and for Group H by the blue line. The shaded areas represent the 90% interquartile range.

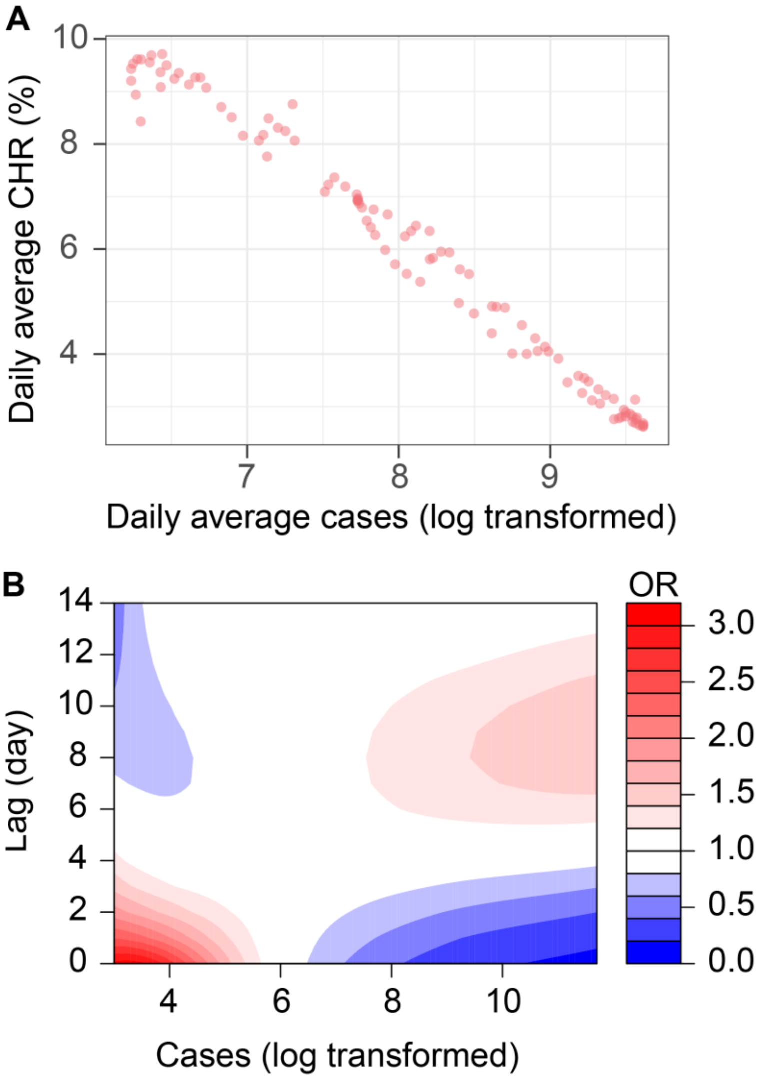


**Figure S13.** Relationship between COVID-19 cases and CHR. (A) Scatter plot of daily average new cases (lag transformed) and daily average CHR at the country level. (B) Lag-response relationships between the number of cases and CHR at different time lags, with redder colors representing higher OR values. A reference value of 400 daily cases was used.

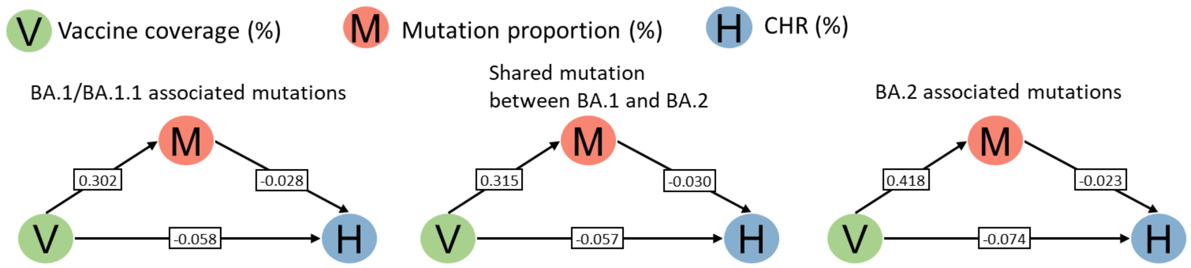


**Figure S14.** Mediation diagram of the alternative SEM model with three variables: V (vaccination coverage, independent), M (mutation proportion, mediator), and H (CHR, dependent). Arrows represent estimated coefficients that indicate the strength and direction of relationships between these variables. For example, positive coefficients between V and M indicate that higher vaccination coverage is directly associated with a greater proportion of mutations, as evidenced by the following coefficients: 0.302, 95% CI: [0.161, 0.443], 0.315, 95% CI: [0.183,0.446], and 0.418, 95% CI: [0.361, 0.475]. Conversely, negative coefficients between M and H indicate an inverse relationship between the proportion of mutations and CHR, as evidenced by the following coefficients: -0.028, 95% CI: [-0.030, -0.026], -0.030, 95% CI: [-0.032, -0.028], and -0.023,95% CI: [-0.027, -0.019]. All coefficients were statistically significant at $p < 0.001$.

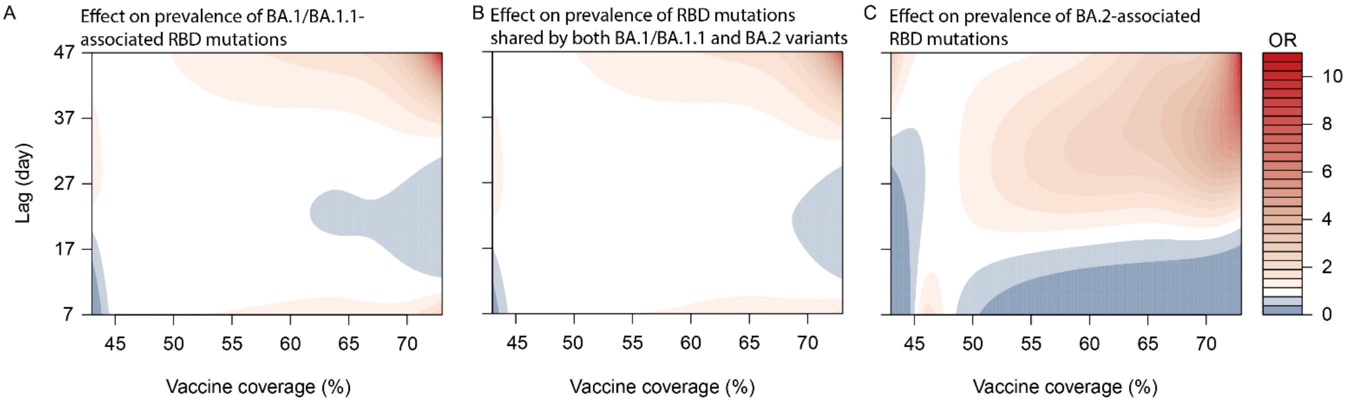


**Figure S15.** The lag-response relationship between vaccination coverage and proportion of Omicron mutations in the sensitivity analysis using lower vaccine effectiveness (i.e., 80%) compared to the main result of 91% effectiveness.

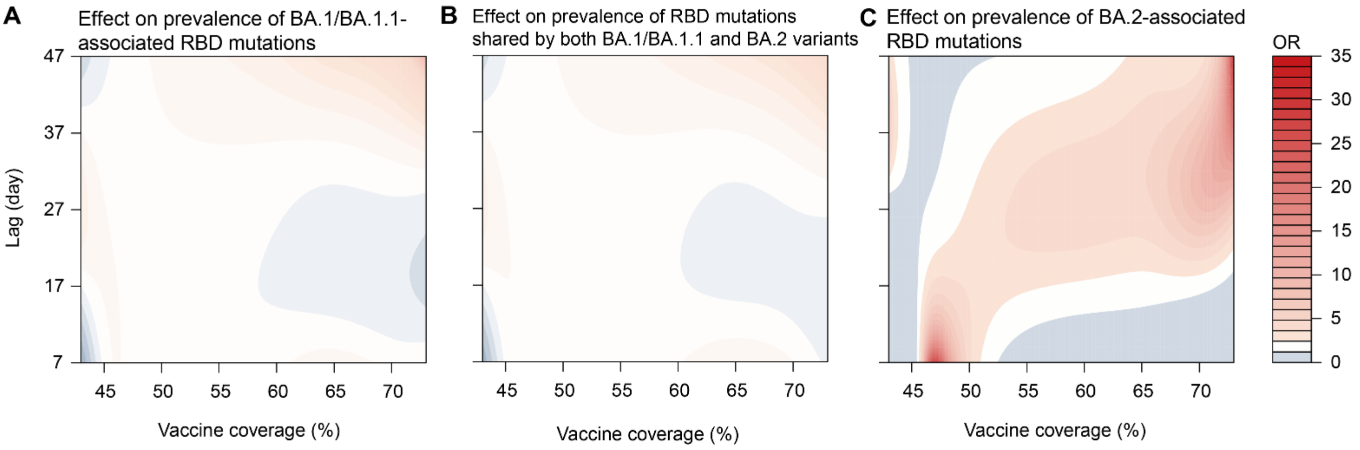


**Figure S16.** The lag-response relationship between vaccination coverage and proportion of Omicron mutations considering pre-immunity effect in the sensitivity analysis. The effect of natural infection in the previous wave was incorporated by adding the cumulative incidence during the Delta wave to the vaccination coverage, representing total population immunity.

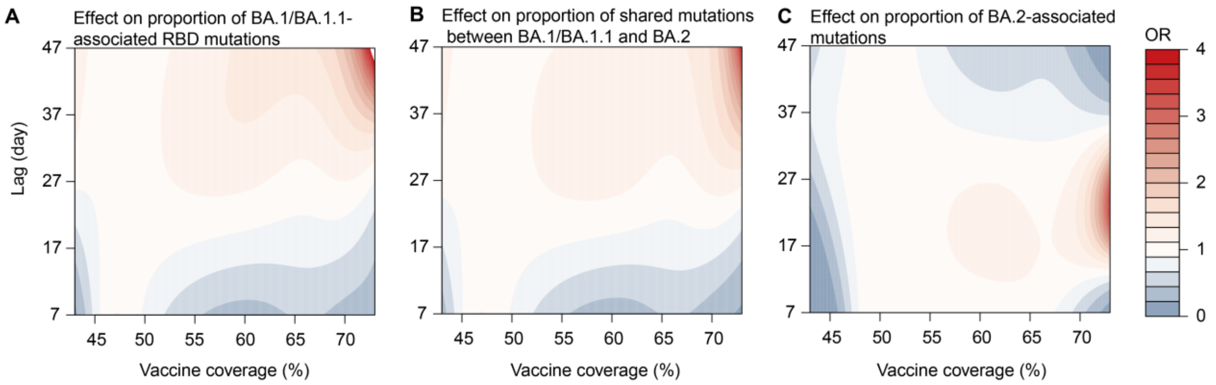


**Figure S17.** The lag-response relationship between vaccination coverage and proportion of Omicron mutations using the alternative *Intermediate model*.

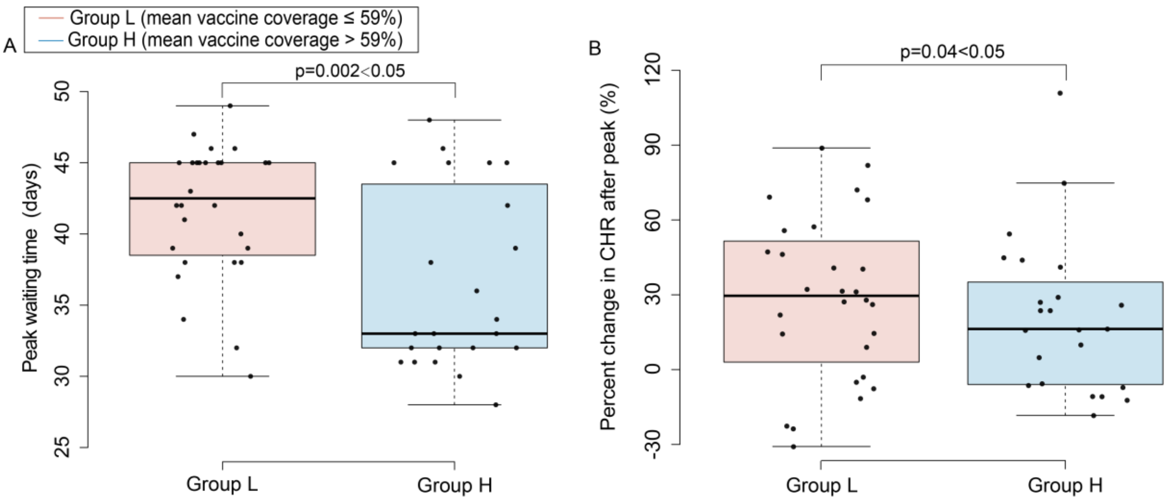


**Figure S18.** Box plot of the peak waiting time (A) and percent change in the daily average CHR after the peak (B) for states in Group L and Group H. The waiting time is defined as the time interval between the study start date of 11 December 2022 and the peak date of each state. The percent change in the daily average CHR after the peak is compared to that before the peak.

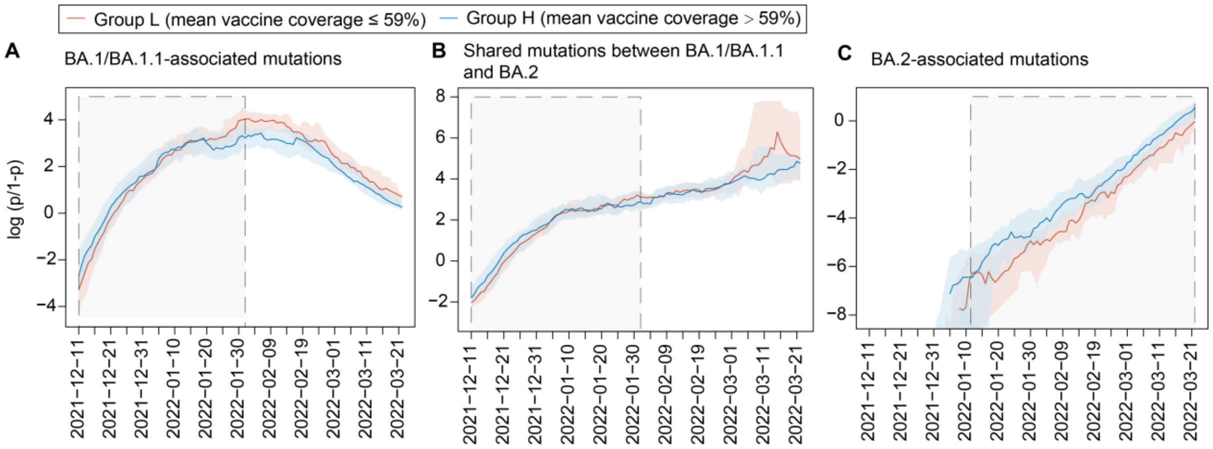


**Figure S19.** Logit transportation of proportion of BA.1/BA.1.1-associated mutations, shared mutations between BA.1/BA.1.1 and BA.2 subvariants, and BA.2-associated mutations in Group L (with low vaccination coverage) and Group H (with high vaccination coverage) states. The grey-shaded areas represented the period during which the proportion of the mutation grew.

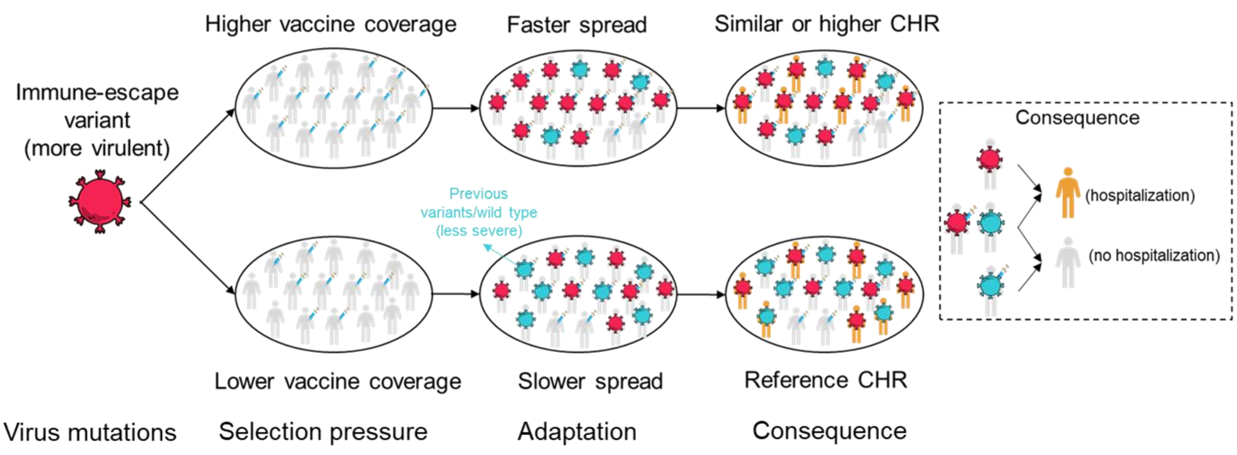


**Figure S20 The impact of vaccination on the transmission advantage of new immune-escape variant and its subsequent effect on the overall CHR amidst increased virulence**. High vaccination coverage might create a selection pressure that accelerates the rapid spread and adaptation of new immune-escape variants. If the emerging variants are more severe than their predecessors (e.g., when vaccination fails to prevent hospitalization in half of the individuals infected with the new variant), high-vaccine-coverage regions might have similar or higher CHR compared to regions with lower vaccination coverage.

#### 3. Supplementary Tables

**Table S1.** Summary statistics of daily reported cases, hospital admissions, and vaccination coverage in the 50 states of the US and the District of Columbia.

| States | Study period | Daily confirmed cases# | Daily hospital admissions# | Vaccination coverage (%)# | Group |
|---|---|---|---|---|---|
| **Alabama (AL)** | 11 Dec,21–22 Mar,22 | 4335 (857,8490) | 181 (57,308) | 46 (45,48) | Group L |
| **Alaska (AK)** | 11 Dec,21–22 Mar,22 | 871 (235,1506) | 12 (7,16) | 55 (53,58) | Group L |
| **Arizona (AZ)** | 11 Dec,21–22 Mar,22 | 6804 (1807,11272) | 294 (177,408) | 55 (54,57) | Group L |
| **Arkansas (AR)** | 11 Dec,21–22 Mar,22 | 2891 (707,4492) | 125 (61,190) | 50 (49,51) | Group L |
| **California (CA)** | 11 Dec,21–22 Mar,22 | 38605 (6603,65818) | 976 (391,1566) | 65 (63,66) | Group H |
| **Colorado (CO)** | 11 Dec,21–22 Mar,22 | 4683 (781,8474) | 168 (63,259) | 64 (63,65) | Group H |
| **Connecticut (CT)** | 11 Dec,21–22 Mar,22 | 2911 (466,4441) | 117 (42,177) | 72 (70,73) | Group H |
| **Delaware (DE)** | 11 Dec,21–22 Mar,22 | 1001 (134,1623) | 41 (14,60) | 62 (61,64) | Group H |
| **District of Columbia (DC)** | 11 Dec,21–22 Mar,22 | 677 (104,1168) | 33 (11,48) | 65 (63,67) | Group H |
| **Florida (FL)** | 11 Dec,21–22 Mar,22 | 20616 (2686,32182) | 889 (264,1476) | 61 (60,62) | Group H |
| **Georgia (GA)** | 11 Dec,21–22 Mar,22 | 7932 (1463,16622) | 373 (140,651) | 50 (48,51) | Group L |
| **Hawaii (HI)** | 11 Dec,21–22 Mar,22 | 1473 (217,2560) | 28 (12,44) | 68 (59,73) | Group H |
| **Idaho (ID)** | 11 Dec,21–22 Mar,22 | 1293 (572,1817) | 39 (22,58) | 48 (44,50) | Group L |
| **Illinois (IL)** | 11 Dec,21–22 Mar,22 | 11708 (1910,19797) | 422 (149,639) | 62 (61,64) | Group H |
| **Indiana (IN)** | 11 Dec,21–22 Mar,22 | 5437 (893,9179) | 265 (97,379) | 50 (49,51) | Group L |
| **Iowa (IA)** | 11 Dec,21–22 Mar,22 | 2084 (509,3205) | 92 (42,130) | 57 (56,58) | Group L |
| **Kansas (KS)** | 11 Dec,21–22 Mar,22 | 3046 (834,5458) | 110 (73,152) | 55 (54,57) | Group L |
| **Kentucky (KY)** | 11 Dec,21–22 Mar,22 | 4899 (1810,8352) | 166 (120,228) | 52 (51,53) | Group L |
| **Louisiana (LA)** | 11 Dec,21–22 Mar,22 | 3853 (506,6464) | 147 (42,257) | 49 (48,50) | Group L |
| **Maine (ME)** | 11 Dec,21–22 Mar,22 | 985 (403,1491) | 28 (15,37) | 73 (72,74) | Group H |
| **Maryland (MD)** | 11 Dec,21–22 Mar,22 | 4044 (637,6742) | 185 (58,276) | 68 (66,70) | Group H |
| **Massachusetts (MA)** | 11 Dec,21–22 Mar,22 | 7200 (1226,11505) | 183 (61,289) | 72 (70,73) | Group H |
| **Michigan (MI)** | 11 Dec,21–22 Mar,22 | 8189 (2063,13938) | 334 (159,489) | 55 (54,56) | Group L |
| **Minnesota (MN)** | 11 Dec,21–22 Mar,22 | 4738 (1276,6936) | 126 (62,173) | 63 (62,65) | Group H |
| **Mississippi (MS)** | 11 Dec,21–22 Mar,22 | 2605 (435,4885) | 87 (25,161) | 47 (46,48) | Group L |
| **Missouri (MO)** | 11 Dec,21–22 Mar,22 | 4693 (1038,7908) | 236 (125,356) | 51 (50,52) | Group L |
| **Montana (MT)** | 11 Dec,21–22 Mar,22 | 759 (159,1316) | 40 (20,62) | 52 (51,53) | Group L |
| **Nebraska (NE)** | 11 Dec,21–22 Mar,22 | 1385 (182,2210) | 53 (35,72) | 58 (57,59) | Group L |
| **Nevada (NV)** | 11 Dec,21–22 Mar,22 | 2090 (575,3508) | 107 (52,164) | 55 (53,56) | Group L |
| **New Hampshire (NH)** | 11 Dec,21–22 Mar,22 | 1285 (581,1663) | 35 (16,51) | 63 (62,65) | Group H |
| **New Jersey (NJ)** | 11 Dec,21–22 Mar,22 | 8886 (1416,13785) | 335 (101,526) | 68 (66,70) | Group H |
| **New Mexico (NM)** | 11 Dec,21–22 Mar,22 | 1874 (513,2741) | 62 (37,84) | 64 (63,66) | Group H |
| **New York (NY)** | 11 Dec,21–22 Mar,22 | 10087 (1536,13855) | 683 (223,1094) | 69 (68,71) | Group H |
| **North Carolina (NC)** | 11 Dec,21–22 Mar,22 | 10473 (2468,18254) | 23 (16,30) | 54 (53,55) | Group L |
| **North Dakota (ND)** | 11 Dec,21–22 Mar,22 | 723 (158,1162) | 23 (16,30) | 51 (50,52) | Group L |
| **Ohio (OH)** | 11 Dec,21–22 Mar,22 | 8984 (1300,14511) | 509 (173,752) | 54 (53,55) | Group L |
| **Oklahoma (OK)** | 11 Dec,21–22 Mar,22 | 3487 (861,5459) | 195 (92,301) | 52 (51,53) | Group L |
| **Oregon (OR)** | 11 Dec,21–22 Mar,22 | 2980 (729,5410) | 73 (40,102) | 64 (63,65) | Group H |
| **Pennsylvania (PA)** | 11 Dec,21–22 Mar,22 | 9618 (2113,14163) | 453 (183,668) | 61 (60,63) | Group H |
| **Rhode Island (RI)** | 11 Dec,21–22 Mar,22 | 1494 (212,2320) | 26 (8,40) | 74 (72,76) | Group H |
| **South Carolina (SC)** | 11 Dec,21–22 Mar,22 | 5294 (1017,9178) | 175 (67,285) | 52 (50,53) | Group L |
| **South Dakota (SD)** | 11 Dec,21–22 Mar,22 | 658 (134,1124) | 33 (18,44) | 55 (54,56) | Group L |
| **Tennessee (TN)** | 11 Dec,21–22 Mar,22 | 6611 (1752,12161) | 218 (117,352) | 50 (49,51) | Group L |
| **Texas (TX)** | 11 Dec,21–22 Mar,22 | 21553 (4533,36940) | 987 (409,1648) | 55 (54,57) | Group L |
| **Utah (UT)** | 11 Dec,21–22 Mar,22 | 3140 (603,5019) | 86 (55,127) | 58 (56,60) | Group L |
| **Vermont (VT)** | 11 Dec,21–22 Mar,22 | 558 (213,697) | 12 (9,16) | 74 (73,76) | Group H |
| **Virginia (VA)** | 11 Dec,21–22 Mar,22 | 6600 (1883,10756) | 238 (122,374) | 66 (64,68) | Group H |
| **Washington (WA)** | 11 Dec,21–22 Mar,22 | 6467 (1641,11744) | 138 (70,207) | 66 (64,67) | Group H |
| **West Virginia (WV)** | 11 Dec,21–22 Mar,22 | 1902 (843,3276) | 81 (61,108) | 53 (53,54) | Group L |
| **Wisconsin (WI)** | 11 Dec,21–22 Mar,22 | 5603 (895,7260) | 208 (91,294) | 60 (59,61) | Group H |
| **Wyoming (WY)** | 11 Dec,21–22 Mar,22 | 427 (97,746) | 14 (9,20) | 47 (45,48) | Group L |

### The mean values (interquartile range) of daily confirmed cases, hospital admissions, vaccination coverage in 50 US states and the District of Columbia

**Table S2.** Summary statistics of daily hospital capacity and weather conditions in the 50 states of the US and the District of Columbia.

| States | Date period | Hospital capacity # | Daily Temperature (°C)# | Daily Relative humidity (%)# |
|---|---|---|---|---|
| **Alabama (AL)** | 11 Dec,21–22 Mar,22 | 14991 (14899,15151) | 10 (7,13) | 71 (64,79) |
| **Alaska (AK)** | 11 Dec,21–22 Mar,22 | 1582 (1534,1620) | -9 (-12,-7) | 78 (76,81) |
| **Arizona (AZ)** | 11 Dec,21–22 Mar,22 | 16489 (16016,16983) | 9 (7,10) | 40 (28,49) |
| **Arkansas (AR)** | 11 Dec,21–22 Mar,22 | 8675 (8010,9098) | 7 (4,10) | 66 (60,73) |
| **California (CA)** | 11 Dec,21–22 Mar,22 | 64721 (63459,65954) | 11 (9,12) | 55 (45,67) |
| **Colorado (CO)** | 11 Dec,21–22 Mar,22 | 11538 (11113,11907) | -3 (-5,-1) | 53 (48,59) |
| **Connecticut (CT)** | 11 Dec,21–22 Mar,22 | 7802 (7698,7852) | 1 (-2,4) | 63 (58,67) |
| **Delaware (DE)** | 11 Dec,21–22 Mar,22 | 3017 (2899,3397) | 5 (1,8) | 65 (59,67) |
| **District of Columbia (DC)** | 11 Dec,21–22 Mar,22 | 3377 (3354,3411) | 5 (1,8) | 53 (49,55) |
| **Florida (FL)** | 11 Dec,21–22 Mar,22 | 57878 (56562,59203) | 18 (16,21) | 71 (68,76) |
| **Georgia (GA)** | 11 Dec,21–22 Mar,22 | 24693 (22101,23171) | 11 (8,14) | 68 (62,75) |
| **Hawaii (HI)** | 11 Dec,21–22 Mar,22 | 2562 (2523,2596) | 21 (21,22) | 75 (71,78) |
| **Idaho (ID)** | 11 Dec,21–22 Mar,22 | 3296 (3048,3326) | -3 (-5,-1) | 75 (71,79) |
| **Illinois (IL)** | 11 Dec,21–22 Mar,22 | 29702 (28658,30414) | 0 (-3,4) | 75 (71,79) |
| **Indiana (IN)** | 11 Dec,21–22 Mar,22 | 16022 (15475,16444) | 0 (-3,4) | 74 (70,76) |
| **Iowa (IA)** | 11 Dec,21–22 Mar,22 | 7596 (7362,7524) | -4 (-8,0) | 71 (67,74) |
| **Kansas (KS)** | 11 Dec,21–22 Mar,22 | 8489 (8348,8723) | 1 (-2,4) | 52 (48,56) |
| **Kentucky (KY)** | 11 Dec,21–22 Mar,22 | 11495 (10767,11957) | 5 (1,8) | 68 (62,73) |
| **Louisiana (LA)** | 11 Dec,21–22 Mar,22 | 12919 (12858,13009) | 13 (11,16) | 69 (63,75) |
| **Maine (ME)** | 11 Dec,21–22 Mar,22 | 3156 (3046,3252) | -5 (-8,-2) | 68 (63,72) |
| **Maryland (MD)** | 11 Dec,21–22 Mar,22 | 10504 (9592,11358) | 5 (1,7) | 64 (59,66) |
| **Massachusetts (MA)** | 11 Dec,21–22 Mar,22 | 17776 (16717,18472) | 1 (-2,4) | 64 (59,69) |
| **Michigan (MI)** | 11 Dec,21–22 Mar,22 | 22067 (21679,22349) | -4 (-7,-1) | 74 (71,76) |
| **Minnesota (MN)** | 11 Dec,21–22 Mar,22 | 9574 (9519,9705) | -10 (-14,-7) | 73 (70,76) |
| **Mississippi (MS)** | 11 Dec,21–22 Mar,22 | 8573 (8198,8813) | 10 (7,13) | 71 (64,79) |
| **Missouri (MO)** | 11 Dec,21–22 Mar,22 | 16549 (15520,17154) | 3 (-1,7) | 63 (58,69) |
| **Montana (MT)** | 11 Dec,21–22 Mar,22 | 2879 (2749,2999) | -4 (-7,0) | 62 (58,67) |
| **Nebraska (NE)** | 11 Dec,21–22 Mar,22 | 4471 (4300,4618) | -2 (-4,1) | 56 (51,61) |
| **Nevada (NV)** | 11 Dec,21–22 Mar,22 | 8321 (8281,8527) | 2 (0,4) | 50 (40,59) |
| **New Hampshire (NH)** | 11 Dec,21–22 Mar,22 | 2890 (2810,2933) | -3 (-6,0) | 66 (59,71) |
| **New Jersey (NJ)** | 11 Dec,21–22 Mar,22 | 23015 (21702,24287) | 3 (0,6) | 65 (59,67) |
| **New Mexico (NM)** | 11 Dec,21–22 Mar,22 | 4132 (4037,4307) | 3 (2,5) | 40 (33,47) |
| **New York (NY)** | 11 Dec,21–22 Mar,22 | 46580 (44533,48506) | -1 (-5,2) | 67 (63,69) |
| **North Carolina (NC)** | 11 Dec,21–22 Mar,22 | 22036 (21802,23319) | 9 (6,11) | 66 (61,71) |
| **North Dakota (ND)** | 11 Dec,21–22 Mar,22 | 2172 (1994,2288) | -11 (-14,-8) | 78 (76,81) |
| **Ohio (OH)** | 11 Dec,21–22 Mar,22 | 29259 (28610,30037) | 1 (-3,5) | 69 (66,72) |
| **Oklahoma (OK)** | 11 Dec,21–22 Mar,22 | 9650 (9206,10164) | 6 (2,9) | 53 (49,59) |
| **Oregon (OR)** | 11 Dec,21–22 Mar,22 | 6989 (6922,7077) | 3 (1,4) | 74 (68,81) |
| **Pennsylvania (PA)** | 11 Dec,21–22 Mar,22 | 30270 (29385,31095) | 1 (-3,4) | 65 (60,67) |
| **Rhode Island (RI)** | 11 Dec,21–22 Mar,22 | 2281 (2259,2347) | 2 (-1,5) | 68 (62,74) |
| **South Carolina (SC)** | 11 Dec,21–22 Mar,22 | 11429 (10942,11762) | 11 (8,14) | 68 (62,74) |
| **South Dakota (SD)** | 11 Dec,21–22 Mar,22 | 2499 (2414,2718) | -6 (-7,-3) | 66 (61,70) |
| **Tennessee (TN)** | 11 Dec,21–22 Mar,22 | 18173 (17013,19368) | 7 (3,10) | 68 (60,75) |
| **Texas (TX)** | 11 Dec,21–22 Mar,22 | 65111 (61516,67908) | 12 (9,15) | 55 (49,61) |
| **Utah (UT)** | 11 Dec,21–22 Mar,22 | 5926 (5576,6210) | 0 (-3,1) | 60 (52,68) |
| **Vermont (VT)** | 11 Dec,21–22 Mar,22 | 1236 (1229,1245) | -5 (-8,-1) | 69 (65,72) |
| **Virginia (VA)** | 11 Dec,21–22 Mar,22 | 17547 (17232,17800) | 6 (2,9) | 63 (58,68) |
| **Washington (WA)** | 11 Dec,21–22 Mar,22 | 11871 (11271,12404) | 3 (2,5) | 80 (77,85) |
| **West Virginia (WV)** | 11 Dec,21–22 Mar,22 | 5717 (5663,5771) | 3 (-1,6) | 66 (61,71) |
| **Wisconsin (WI)** | 11 Dec,21–22 Mar,22 | 12469 (12107,12825) | -6 (-10,-4) | 68 (65,71) |
| **Wyoming (WY)** | 11 Dec,21–22 Mar,22 | 1704 (1623,1743) | -4 (-6,-1) | 56 (52,61) |

### The mean values (interquartile range) of hospital capacity, temperature, and relative humidity in 50 US states and the District of Columbia

**Table S3.** The number of SARS-Cov-2 virus sequence samples for each state.

| States | Date period | Number of sequences |
|---|---|---|
| **Alabama (AL)** | 11 Dec,21–22 Mar,22 | 14587 |
| **Alaska (AK)** | 11 Dec,21–22 Mar,22 | 5381 |
| **Arizona (AZ)** | 11 Dec,21–22 Mar,22 | 64147 |
| **Arkansas (AR)** | 11 Dec,21–22 Mar,22 | 8872 |
| **California (CA)** | 11 Dec,21–22 Mar,22 | 410799 |
| **Colorado (CO)** | 11 Dec,21–22 Mar,22 | 123740 |
| **Connecticut (CT)** | 11 Dec,21–22 Mar,22 | 33323 |
| **Delaware (DE)** | 11 Dec,21–22 Mar,22 | 10307 |
| **District of Columbia (DC)** | 11 Dec,21–22 Mar,22 | 6401 |
| **Florida (FL)** | 11 Dec,21–22 Mar,22 | 121743 |
| **Georgia (GA)** | 11 Dec,21–22 Mar,22 | 48511 |
| **Hawaii (HI)** | 11 Dec,21–22 Mar,22 | 11218 |
| **Idaho (ID)** | 11 Dec,21–22 Mar,22 | 20463 |
| **Illinois (IL)** | 11 Dec,21–22 Mar,22 | 63173 |
| **Indiana (IN)** | 11 Dec,21–22 Mar,22 | 29496 |
| **Iowa (IA)** | 11 Dec,21–22 Mar,22 | 10523 |
| **Kansas (KS)** | 11 Dec,21–22 Mar,22 | 12978 |
| **Kentucky (KY)** | 11 Dec,21–22 Mar,22 | 19402 |
| **Louisiana (LA)** | 11 Dec,21–22 Mar,22 | 22203 |
| **Maine (ME)** | 11 Dec,21–22 Mar,22 | 14590 |
| **Maryland (MD)** | 11 Dec,21–22 Mar,22 | 40741 |
| **Massachusetts (MA)** | 11 Dec,21–22 Mar,22 | 95638 |
| **Michigan (MI)** | 11 Dec,21–22 Mar,22 | 59927 |
| **Minnesota (MN)** | 11 Dec,21–22 Mar,22 | 79124 |
| **Mississippi (MS)** | 11 Dec,21–22 Mar,22 | 9433 |
| **Missouri (MO)** | 11 Dec,21–22 Mar,22 | 19127 |
| **Montana (MT)** | 11 Dec,21–22 Mar,22 | 11839 |
| **Nebraska (NE)** | 11 Dec,21–22 Mar,22 | 12137 |
| **Nevada (NV)** | 11 Dec,21–22 Mar,22 | 21498 |
| **New Hampshire (NH)** | 11 Dec,21–22 Mar,22 | 8747 |
| **New Jersey (NJ)** | 11 Dec,21–22 Mar,22 | 52133 |
| **New Mexico (NM)** | 11 Dec,21–22 Mar,22 | 27476 |
| **New York (NY)** | 11 Dec,21–22 Mar,22 | 140544 |
| **North Carolina (NC)** | 11 Dec,21–22 Mar,22 | 61839 |
| **North Dakota (ND)** | 11 Dec,21–22 Mar,22 | 8527 |
| **Ohio (OH)** | 11 Dec,21–22 Mar,22 | 39222 |
| **Oklahoma (OK)** | 11 Dec,21–22 Mar,22 | 4972 |
| **Oregon (OR)** | 11 Dec,21–22 Mar,22 | 37467 |
| **Pennsylvania (PA)** | 11 Dec,21–22 Mar,22 | 50428 |
| **Rhode Island (RI)** | 11 Dec,21–22 Mar,22 | 13042 |
| **South Carolina (SC)** | 11 Dec,21–22 Mar,22 | 19498 |
| **South Dakota (SD)** | 11 Dec,21–22 Mar,22 | 4156 |
| **Tennessee (TN)** | 11 Dec,21–22 Mar,22 | 51853 |
| **Texas (TX)** | 11 Dec,21–22 Mar,22 | 171588 |
| **Utah (UT)** | 11 Dec,21–22 Mar,22 | 79351 |
| **Vermont (VT)** | 11 Dec,21–22 Mar,22 | 12686 |
| **Virginia (VA)** | 11 Dec,21–22 Mar,22 | 34310 |
| **Washington (WA)** | 11 Dec,21–22 Mar,22 | 76114 |
| **West Virginia (WV)** | 11 Dec,21–22 Mar,22 | 24619 |
| **Wisconsin (WI)** | 11 Dec,21–22 Mar,22 | 42699 |
| **Wyoming (WY)** | 11 Dec,21–22 Mar,22 | 20539 |

**Table S4.** Model Selection Based on the Akaike Information Criterion (AIC). Model 1 is the baseline model and Model 5 is the best-fitting model.

| | **Model** | **AIC** |
|---|---|---|
| 1 | $\log\left(\frac{CHR_{i,t}}{1-CHR_{i,t}}\right) = \alpha_i + aVac_{i,t-7}$ | 305613.0 |
| 2 | $\log\left(\frac{CHR_{i,t}}{1-CHR_{i,t}}\right) = \alpha_i + aVac_{i,t-7} + \sum_{lag=0}^{14} cb\left(Log(cases)_{i,t}, lag\right)$ | 71562.3 |
| 3 | $\log\left(\frac{CHR_{i,t}}{1-CHR_{i,t}}\right) = \alpha_i + aVac_{i,t-7} + \sum_{lag=0}^{14} cb\left(Log(cases)_{i,t}, lag\right) + \beta_1 Hospital\ capacity_{i,t}$ | 70784.4 |
| 4 | $\log\left(\frac{CHR_{i,t}}{1-CHR_{i,t}}\right) = \alpha_i + aVac_{i,t-7} + \sum_{lag=0}^{14} cb\left(Log(cases)_{i,t}, lag\right) + \beta_1 Hospital\ capacity_{i,t} + \beta_2 Temp_{it,1-14} + \beta_3 RH_{it,1-14}$ | 69100.8 |
| 5 | $\log\left(\frac{CHR_{i,t}}{1-CHR_{i,t}}\right) = \alpha_i + aVac_{i,t-7} + \sum_{lag=0}^{14} cb\left(Log(cases)_{i,t}, lag\right) + \beta_1 Hospital\ capacity_{i,t} + \beta_2 Temp_{it,1-14} + \beta_3 RH_{it,1-14} + I(Holiday_{i,t}) + I(Weekend_{i,t})$ | 69063.3 |

**Table S5.** The effect of factors on CHR after using lower vaccine effectiveness (i.e., 80%) after six months of full vaccination against hospitalization, compared to the effectiveness of 91% used in the main result.

| | **Basic Model** (CHR~vaccine) | | **Mediation Model** (CHR~vaccine+virus mutation) | |
|---|---|---|---|---|
| | **OR (95%CI)** | **P value** | **OR (95%CI)** | **P value** |
| **Vaccine** | | | | |
| Vaccination coverage (25%) | 0.434 (0.420, 0.448) | <0.001 | 0.710 (0.643, 0.786) | <0.001 |
| **Virus mutation** | | | | |
| Mean prevalence of BA.1-specific mutations (20%) | - | - | 0.861 (0.831, 0.892) | <0.001 |
| Mean prevalence of BA.2-specific mutations (20%) | - | - | 0.754 (0.727, 0.782) | <0.001 |
| Mean prevalence of shared mutations between BA.1 and BA.2 (20%) | - | - | 0.996 (0.958, 1.036) | 0.702 |
| **Confounders** | | | | |
| Temperature (5℃) | 1.082 (1.077, 1.086) | <0.001 | 1.086 (1.079, 1.093) | <0.001 |
| Relative humidity (5%) | 1.020 (1.018, 1.022) | <0.001 | 1.013 (1.011, 1.016) | <0.001 |
| Hospital beds (1000) | 1.003 (1.002, 1.004) | <0.001 | 1.003 (1.001, 1.004) | <0.001 |
| Weekend (Yes vs No) | 1.001 (0.997, 1.005) | 0.532 | 1.002 (0.998, 1.007) | 0.295 |
| Holiday (Yes vs No) | 1.032 (1.022, 1.041) | <0.001 | 1.021 (1.011, 1.031) | <0.001 |

**Table S6.** The effect of factors on CHR after adding the cumulative incidence during the Delta wave to the vaccination coverage in the sensitivity analysis.

| | **Basic Model** (CHR~vaccine) | | **Mediation Model** (CHR~vaccine+virus mutation) | |
|---|---|---|---|---|
| | **OR (95%CI)** | **P value** | **OR (95%CI)** | **P value** |
| **Vaccine** | | | | |
| Vaccination coverage (25%) | 0.392 (0.406, 0.379) | <0.001 | 0.804 (0.712, 0.908) | <0.001 |
| **Virus mutation** | | | | |
| Mean prevalence of BA.1-specific mutations (20%) | - | - | 0.935 (0.919, 0.952) | <0.001 |
| Mean prevalence of BA.2-specific mutations (20%) | - | - | 0.868 (0.853, 0.884) | <0.001 |
| Mean prevalence of shared mutations between BA.1 and BA.2 (20%) | - | - | 0.994 (0.975, 1.014) | 0.549 |
| **Confounders** | | | | |
| Temperature (5℃) | 1.092 (1.087, 1.096) | <0.001 | 1.088 (1.081, 1.094) | <0.001 |
| Relative humidity (5%) | 1.017 (1.015, 1.019) | <0.001 | 1.012 (1.010, 1.015) | <0.001 |
| Hospital beds (1000) | 1.003 (1.002, 1.004) | <0.001 | 1.002 (1.001, 1.004) | <0.001 |
| Weekend (Yes vs No) | 1.001 (0.997, 1.006) | 0.532 | 1.003 (0.998, 1.007) | 0.276 |
| Holiday (Yes vs No) | 1.030 (1.021, 1.040) | <0.001 | 1.022 (1.012, 1.031) | <0.001 |
